# Maternal-fetal outcomes in patients with immune mediated inflammatory diseases, with consideration of comorbidities: a retrospective cohort study in a large U.S. healthcare system

**DOI:** 10.1101/2023.08.07.23293726

**Authors:** Yeon Mi Hwang, Qi Wei, Samantha N. Piekos, Bhargav Vemuri, Sevda Molani, Philip Mease, Leroy Hood, Jennifer J. Hadlock

## Abstract

**Background:** Immune-mediated inflammatory diseases (IMIDs) are likely to complicate maternal health. However, literature data on patients with IMIDs undergoing pregnancy is scarce and often overlooks the impact of comorbidities.

**Methods:** We investigated 12 selected IMIDs: psoriasis, inflammatory bowel disease, rheumatoid arthritis, spondyloarthritis, multiple sclerosis, systemic lupus erythematosus, psoriatic arthritis, antiphospholipid syndrome, Sjögren’s syndrome, vasculitis, sarcoidosis, systemic sclerosis. We characterized patients with IMIDs prior to pregnancy (IMIDs group) based on pregnancy/maternal characteristics, comorbidities, and pre-pregnancy/prenatal immunomodulatory medications (IMMs) prescription patterns. We 1:1 propensity score matched the IMIDs cohort with people who had no IMID diagnoses prior to pregnancy (non-IMIDs cohort). Outcome measures were preterm birth (PTB), low birth weight (LBW), small for gestational age (SGA), and cesarean section.

**Findings:** The prevalence rate of pregnancy occurring with people with a previous IMID diagnosis has doubled in the past ten years. We identified 5,784 patients with IMIDs. 17% of the IMIDs group had at least one prenatal IMM prescription. Depending on the type of IMM, from 48% to 70% of the patients taking IMMs before pregnancy continued them throughout pregnancy. Patients with IMIDs had similar but slightly increased risks of PTB (Relative risk (RR)=1·1[1·0, 1·3]), LBW (RR=1·2 [1·0,1·4]), SGA (RR=1·1 [1·0,1·2]), and cesarean section (RR=1·1 [1·1,1·2]) compared to a matched cohort of people without IMIDs. Out of the 12 selected IMIDs, three for PTB, one for LBW, two for SGA, and six for cesarean section had results supporting increased risk.

**Interpretation:** The association between IMIDs and the increased risk of adverse pregnancy outcomes depend on both the nature of the IMID and the presence of comorbidities.

**Funding:** NIH

## Introduction

Immune-mediated inflammatory disorders (IMIDs) are a group of conditions with heterogeneous clinical presentation that share some common pathogenic immune pathways and affect multiple human body organ systems.^1^ IMIDs are generally characterized by organ damage and chronic inflammation, resulting in reduced quality of life, comorbidities, and premature death.^2^ Although each individual IMID has unique epidemiology and pathophysiology, its pathogenesis is primarily attributable to an imbalance in immune cellular activation and inflammatory cytokines.^1^ The underlying causes and mechanisms for the pathogenesis of many IMIDs remains ill-defined, but there have been significant therapeutic advances over the past two decades.^1^ We investigated a partial list of IMIDs in this study: psoriasis (PsO), inflammatory bowel disease (IBD), rheumatoid arthritis (RA), spondyloarthritis (SpA), multiple sclerosis (MS), systemic lupus erythematosus (SLE), psoriatic arthritis (PsA), antiphospholipid syndrome (APS), Sjögren’s syndrome (SjS), vasculitis (Va), sarcoidosis (Sa), systemic sclerosis (SSc) (order based on number of IMIDs diagnosis in our study, see Figure 2C for more detail).

Although the degree of sexual bias varies widely across individual IMIDs, most IMIDs occur more frequently in females than males; 80% of patients with autoimmune diseases are female.^3^. RA, IBD, MS, SLE, APS, SjS and SSc occur 2-3^4^ (RA), 1.5^5^ (IBD), 2^6^ (MS), 7-10^7^ (SLE), 3.5^8^ (APS), 13^9^ (SjS), and 3-8^10^ (SSc) times more often in females than males. Given insufficient understanding of the pathology and mechanisms of IMIDs, the underlying reason for sexual dimorphism in IMIDs is still unknown.

It is particularly important to evaluate the relationship between pregnancy and IMIDs because IMID are often first diagnosed during reproductive age.Pregnancy can ameliorate or exacerbates disease activity, depending on the specific IMID.^11^ Both MS and RA can improve during pregnancy and flare after the delivery.^11^ SLE can induce unpredictable changes in disease activity during pregnancy and is one of the most significant risk factors for adverse pregnancy outcomes.^11–14^ Less common autoimmune rheumatic diseases (e.g. PsA) were also shown to associate with worse outcomes including PTB, SGA.^15^ Recent meta-analysis studies on RA and IBD reported elevated risk of adverse pregnancy outcomes including PTB, LBW, LBW, cesarean section or stillbirth.^16, 17^ Pregnancy itself is a significant perturbation in the maternal immune system; the maternal immune system has to avoid rejecting a semi-allogeneic fetus while remaining immunocompetent.^18^ Because IMIDs can further complicate pregnancy and patient’s health, patients often voluntarily avoid pregnancy. Patients with IBD had significantly higher rate of voluntary childlessness, ranging from 14-18%, compared to 6·8% of the general population.^19^ However, recent years have shown improvements in pregnancy outcomes through substantial progress in diagnosis, and in preconception and prenatal care.^20^

Comorbid conditions, are common in patients with IMIDs, including cardiovascular disease, metabolic and bone disorders and cognitive deficit.^1^ Also, patients with IMIDs have increased incidence of psychiatric disorders including depression, anxiety, and bipolar disorder, compared to geographically-age-, and sex-matched controls.^21^ Despite the high cooccurrence of comorbidities among patients with IMIDs condition, the impact of comorbidities on the relation between IMIDs and pregnancy course is insufficiently examined.

The objective of this study is to retrospectively characterize patients who had one or more diagnoses of IMIDs prior to pregnancy regarding their demographics, pregnancy characteristics, comorbidities, and use of immunomodulatory drugs. We will evaluate the impact of IMIDs on adverse pregnancy outcomes after assessing and addressing any discrepancies in the distribution of covariates associated with adverse pregnancy outcomes between patients with and without IMIDs. We will also conduct disease-specific and sensitivity analyses to examine the contribution of individual IMIDs and comorbidities on the relationship between IMIDs and adverse pregnancy outcomes.

## Method

### Study setting and participants

Providence Health and Services and affiliates (PHSA) is an integrated U.S. community healthcare system that provides care in urban and rural settings across seven states: Alaska, California, Montana, Oregon, New Mexico, Texas, and Washington. We used PHSA electronic health records (EHR) of pregnant people who delivered from January 1, 2013, through December 31, 2022 (n=543,408). Figure 1 describes the cohort selection. We included pregnant people with age between 18 and 45 years (n = 510,488). To reduce surveillance bias, we included people who had continuity of care at PHSA. Continuous enrollment was defined as at least one encounter 180 days prior to conception and one encounter on or after the delivery date. We excluded multifetal gestations and deliveries with gestational age (GA) of less than 20 weeks (n = 516,881).

**Figure 1.**
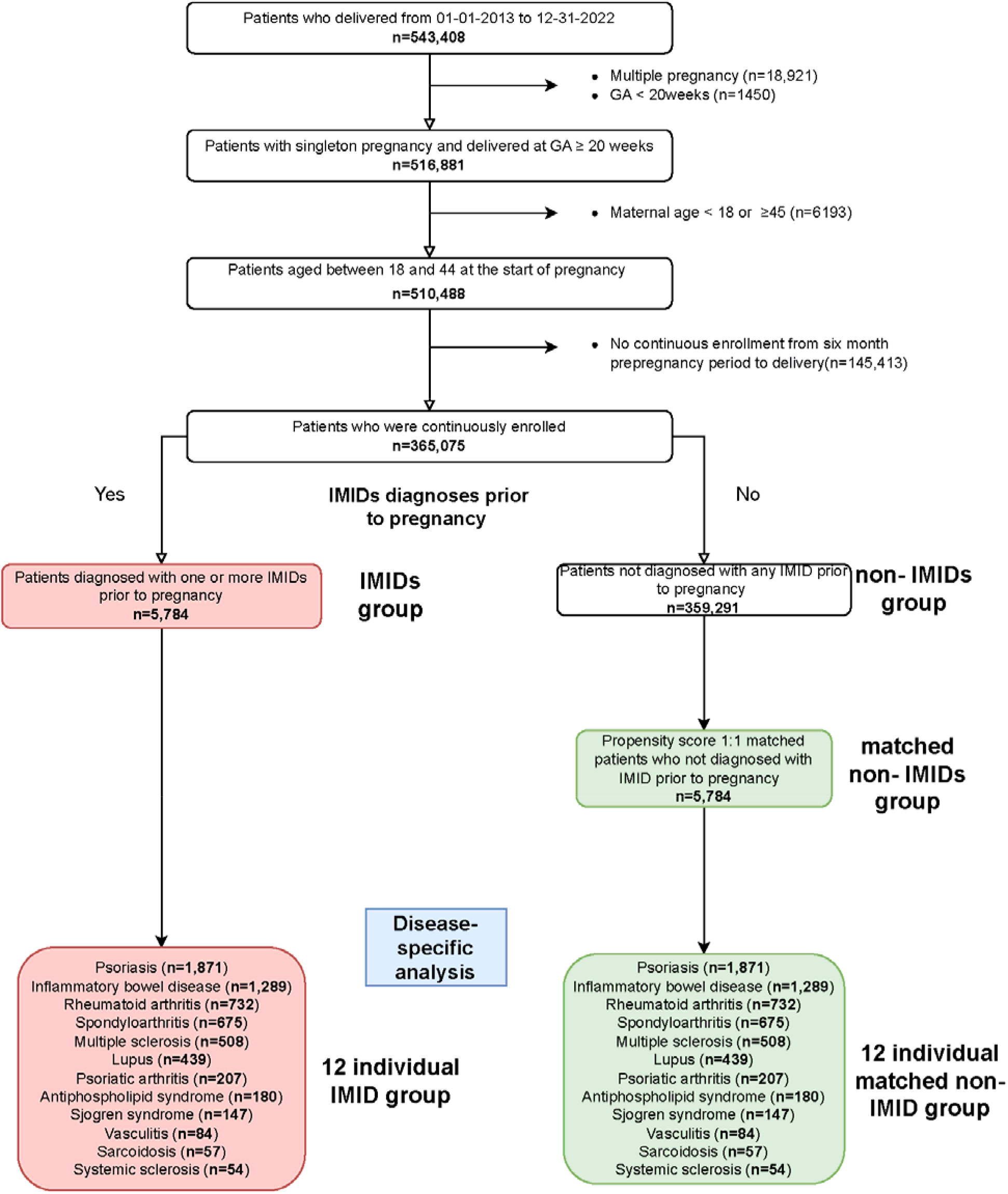
Cohort selection flow chart. GA Gestational Age; IMIDs Immune-mediated Inflammatory Disease IMIDs group was propensity score matched 1:1 on confounding variables to generate the matched non-IMIDs group. Individual IMID groups were propensity score matched 1:1 on pregnancy/maternal characteristics and comorbidities variables to generate corresponding matched control groups.

All procedures were reviewed and approved by the Institutional Review Board at the PHSA through expedited review on 11-04-2020 (study number STUDY2020000196). Consent was waived because disclosure of protected health information for the study involved no more than minimal risk to the privacy of individuals.

### Exposure and outcomes

Our exposure of interest was pre-existing IMIDs conditions prior to pregnancy. The IMIDs group was people who had at least one type of IMIDs before pregnancy (n = 5784). The twelve IMIDs we investigated were PsO, IBD, RA, SpA, MS, SLE, PsA, APS, SjS, Va, Sa, and SSc. SNOMED diagnoses of these IMIDs were clinically reviewed (Table S1). People with no IMIDs diagnosis before pregnancy comprised the control group, the non-IMID group (Figure 1).

Our outcomes of interest were adverse pregnancy outcomes: preterm birth (PTB; birth before 37 weeks of gestational age), low birthweight (LBW; birth of baby weighing less than 2,500g), small for gestational age (SGA; birth of baby weighing less than 10 percentile of babies born in same gestational age), and cesarean section (Table S2).

### Cohort characteristics

We collected information on maternal/pregnancy characteristics, comorbid conditions, and pre-pregnancy/prenatal IMMs prescription pattern (Table S2). The included maternal/pregnancy characteristics are parity, gravidity, preterm history, delivery year, fetal sex, maternal age, pregravid body mass index (BMI) category, self-reported smoking status, self-reported illegal drug use status, self-reported racial group, self-reported ethnic group, insurance, Centers for Disease Control and Prevention social vulnerability index (CDC-SVI), and rural-urban classification (Table S2). CDC-SVI represents the percentile ranking of each census tract on 15 social factors. Social factor themes include socioeconomic status, household composition, race/ethnicity/language, and housing/transportation. CDC-SVI and rural/urban classification were collected based on the census tract patient resided in. Maternal and pregnancy characteristics were collected at the time of the prenatal visit. We included the following comorbidities: urinary tract infection, sexually transmitted disease, obesity, diabetes, asthma, depression, chronic kidney disease, chronic lung disease, pneumonia, sepsis, and hypercoagulability. Comorbidities that were active at the start of pregnancy (last menstrual period; LMP) were collected.

We assessed the pre-pregnancy and prenatal prescription patterns of IMMs. We defined prescription status based on prescription records within the period of interest. We limited our search to records with administration routes of oral, intramuscular, intravenous, subcutaneous, and rectal. The pre-pregnancy study period was 180 days from LMP to LMP. Prenatal medication exposure was further categorized based on their administration status in each trimester. IMMs we investigated were hydroxychloroquine, methotrexate, leflunomide and teriflunomide, 5-ASA, azathioprine, mercaptopurine, mitoxantrone, mycophenolate, calcineurin inhibitors, TNF-α inhibitor, fumarates, interferon, alkylating agent, hydroxyurea, dapsone, cladribine, IL-1 inhibitors, IL-6 inhibitors, IL-12/23 inhibitors, IL-17 inhibitors, IL-12/23 inhibitors, IL-23 inhibitors, abatacept, anti-BLyS, S1P receptor modulator, JAK inhibitor, PDE4 inhibitor, anti-CD20, anti-CD52, budesonide, and systemic glucocorticoids (Table S3).

### Descriptive statistics

We described pregnancy/maternal characteristics, comorbidities, and pre-pregnancy/prenatal IMMs prescription patterns. We calculated the mean and standard deviation for continuous variables and the proportion of categories for binary and multi-class categorical variables. Differences in the distribution of variables between IMID and non-IMID groups were evaluated using the t-test, fisher exact test, and chi-square test for continuous, binary, and multi-class categorical variables. Multiple testing errors were corrected using the Benjamini-Hochberg method.

### Propensity score matching

We generated a matched non-IMIDs group using the propensity score matching method. Propensity score was calculated across covariates using logistic regression (scikit-learn v1.0.2). Covariates include parity, gravidity, preterm history, delivery year, fetal sex, maternal age, pregravid BMI, self-reported smoking status, self-reported illegal drug use status, self-reported racial group, self-reported ethnic group, race, ethnicity, insurance, pregravid BMI, smoking status, illegal drug use status, rural-urban classification, and comorbidities. People in the IMID group were 1:1 matched to the non-IMID group using the K-nearest neighbor algorithm in Euclidean metrics (k=1), which identifies for each IMIDs person the most similar non-IMIDs person across several features in high-dimensional space (scikit-learn v1.0.2).

### Disease-specific analysis and sensitivity analysis

We replicated descriptive statistics and propensity score matching on individual IMIDs. Patients of individual IMID groups were 1:1 matched with non-IMIDs people to generate individual matched control groups. For sensitivity analysis, we conducted propensity score matching without comorbidities to examine the influence of comorbidities on the associations between IMIDs and adverse pregnancy outcomes (Figure 1). Sensitivity analysis assessing the impact of comorbidities on the association was conducted on individual IMID groups.

## Result

### Identification of IMIDs, individual IMID, and matched control cohorts

Our analytic cohort had 365,075 people, of which 5784 were in the IMIDs group and 359,291 were in the non-IMIDs group (Figure 1, continuously enrolled singleton pregnant people). The IMIDs group’s most and least common IMID diagnoses were PsO and SSc respectively (PsO n = 1,871; IBD n = 1,289; RA n = 732; SpA n = 675; MS n = 508; SLE n = 439; PsA n = 207; APS n = 180; SjS n = 147; Va n = 84; Sc n = 57; SSc n = 54). IMIDs people were 1:1 matched to non-IMID people to generate matched non-IMIDs groups (n=5784).

### Characteristics of IMIDs and individual IMID cohorts

Figure 1 and Table 1 describe non-IMIDs, IMIDs, and the subclassification of the IMIDs group into twelve distinct IMID groups. The prevalence rate of pregnancy with prior IMIDs diagnoses increased from 1.1 to 2.0% in the recent ten years (Figure 1). It escalated steadily from 2013 to 2020 and plateaued from 2020 to 2022. Compared to the non-IMIDs group, people in the IMID group (n = 5,784) were more likely to identify as white (73·1% vs. 62·9%, p<0·0001) or non-Hispanic or Latino (86·2% vs. 76·6%, p<0·0001), be between the ages of 30-34 (54·8% vs. 43·1%, P < 0·0001), and have commercial insurance (55·2% vs. 46·3%, p<0·0001). Regarding comorbidities, the IMIDs group had significantly more people with cardiovascular disease (35·0% vs 13·9%, p< 0.0001), depression (22·1% vs 9·1%, p<0.0001), urinary tract infection (18·8% vs. 10·4%, p<0.0001), asthma (12·0% vs 5·2%, p<0.0001), obesity (7·0% vs 3·0%, p<0.0001), sexually transmitted disease (3·5% vs 1·7%, p<0.0001), and diabetes (1·5% vs 0·7%, p<0.0001).

**Table 1.**
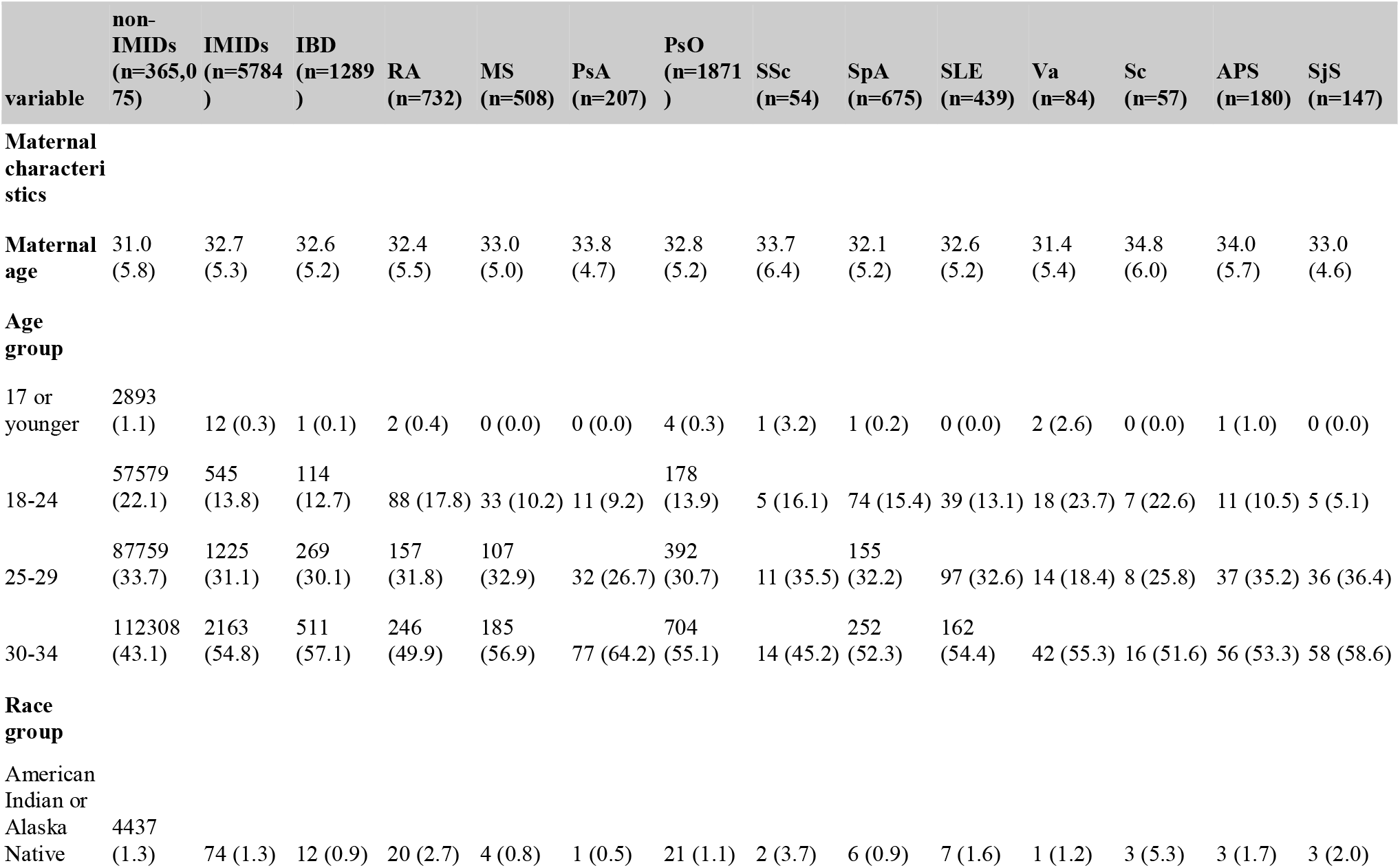
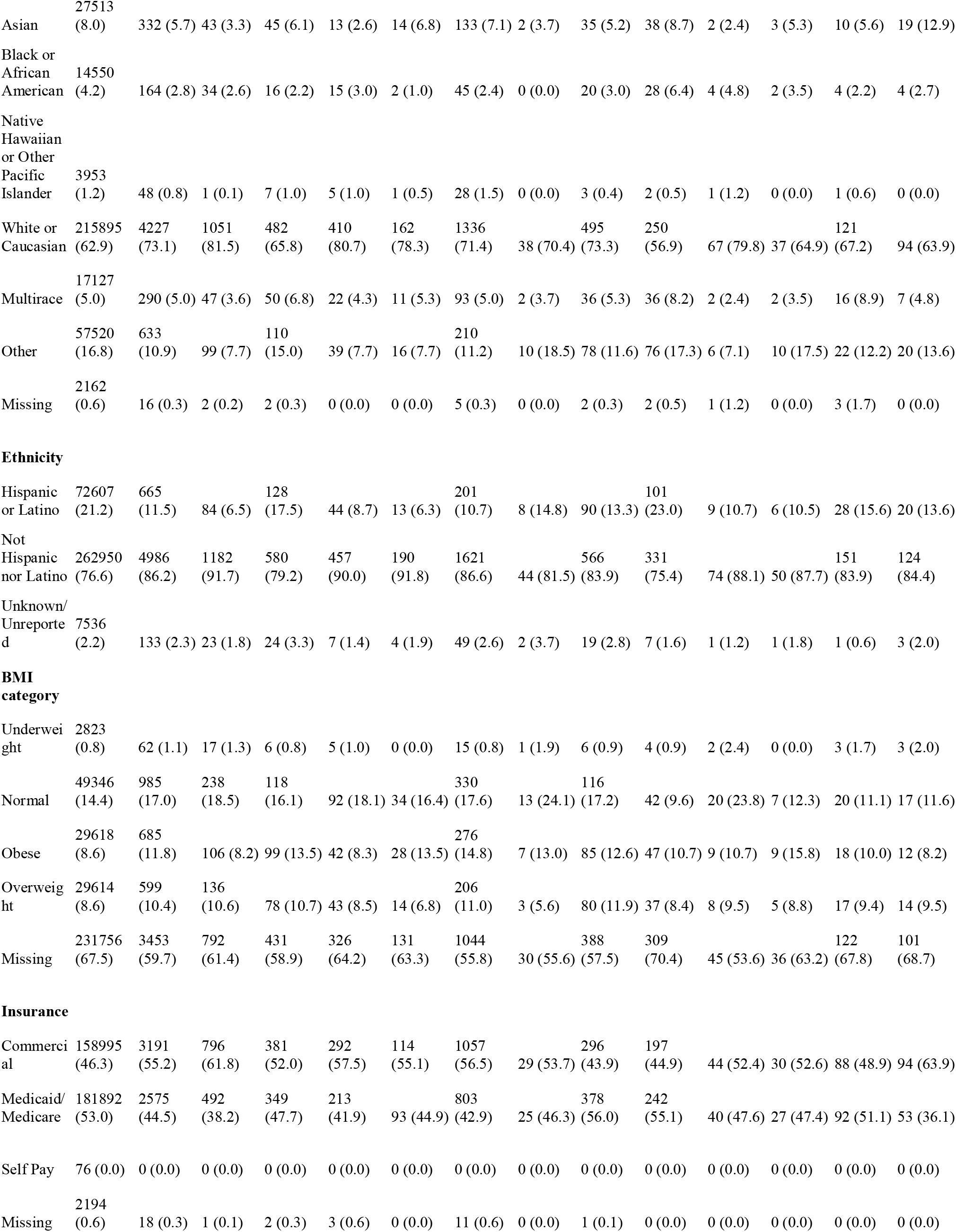
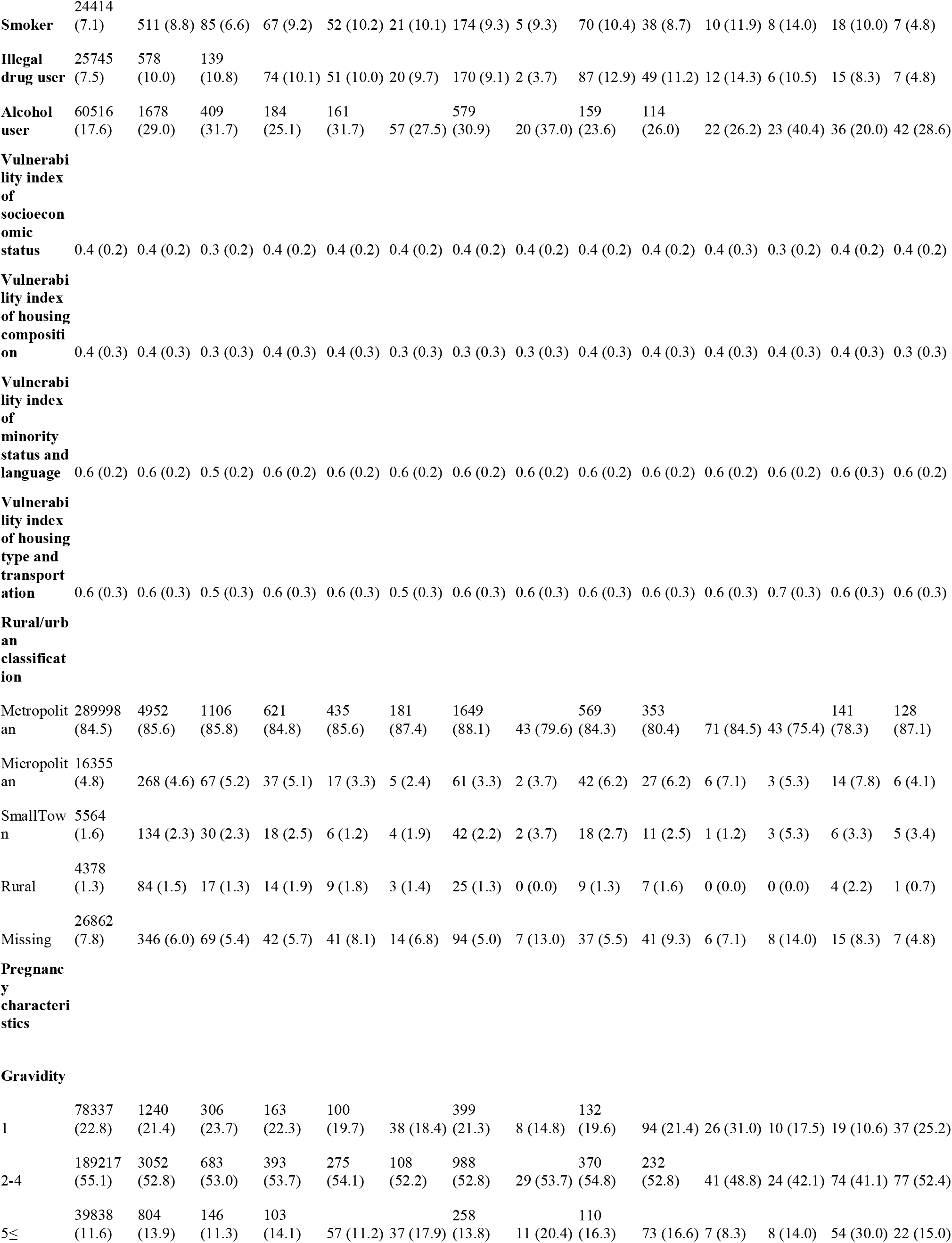
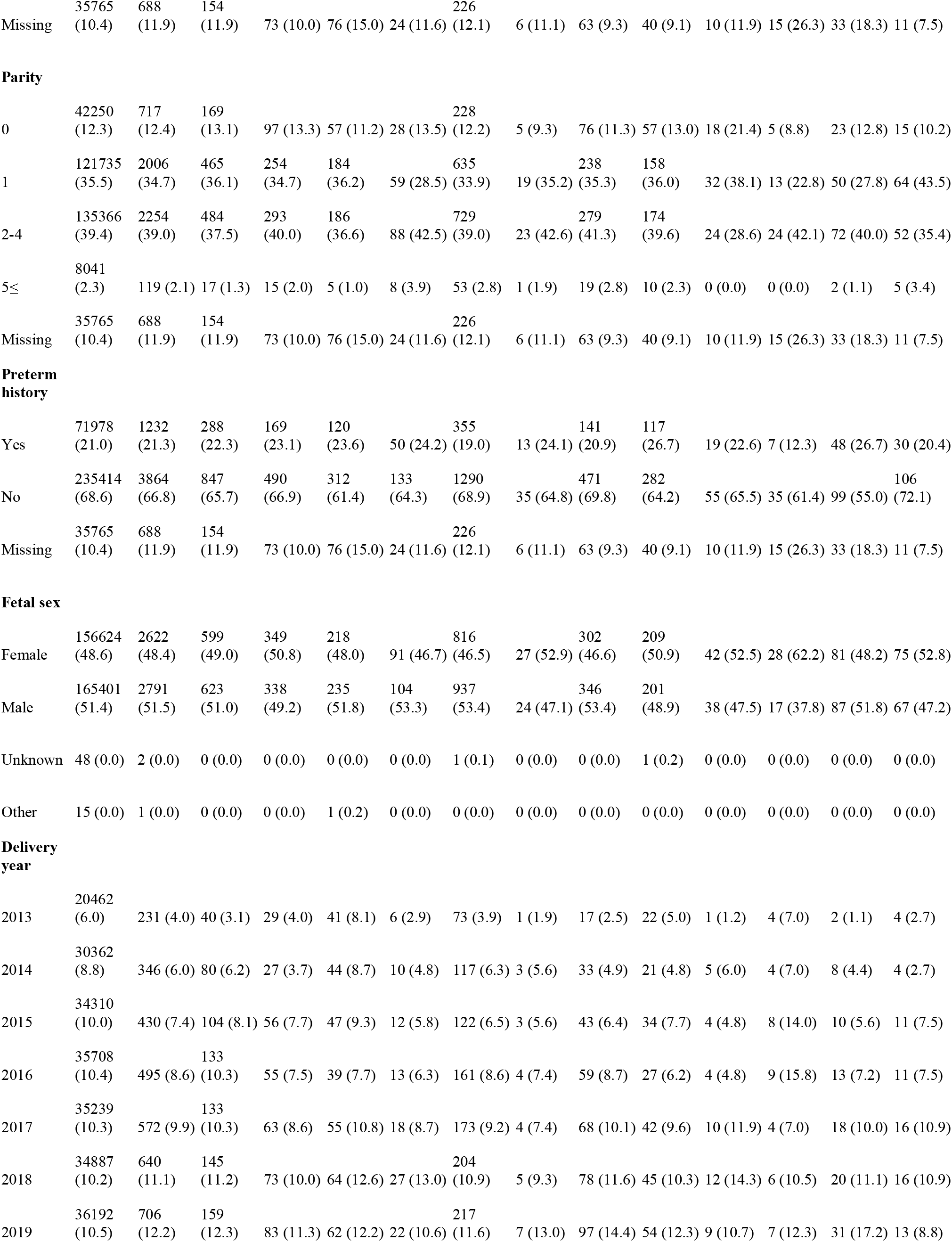
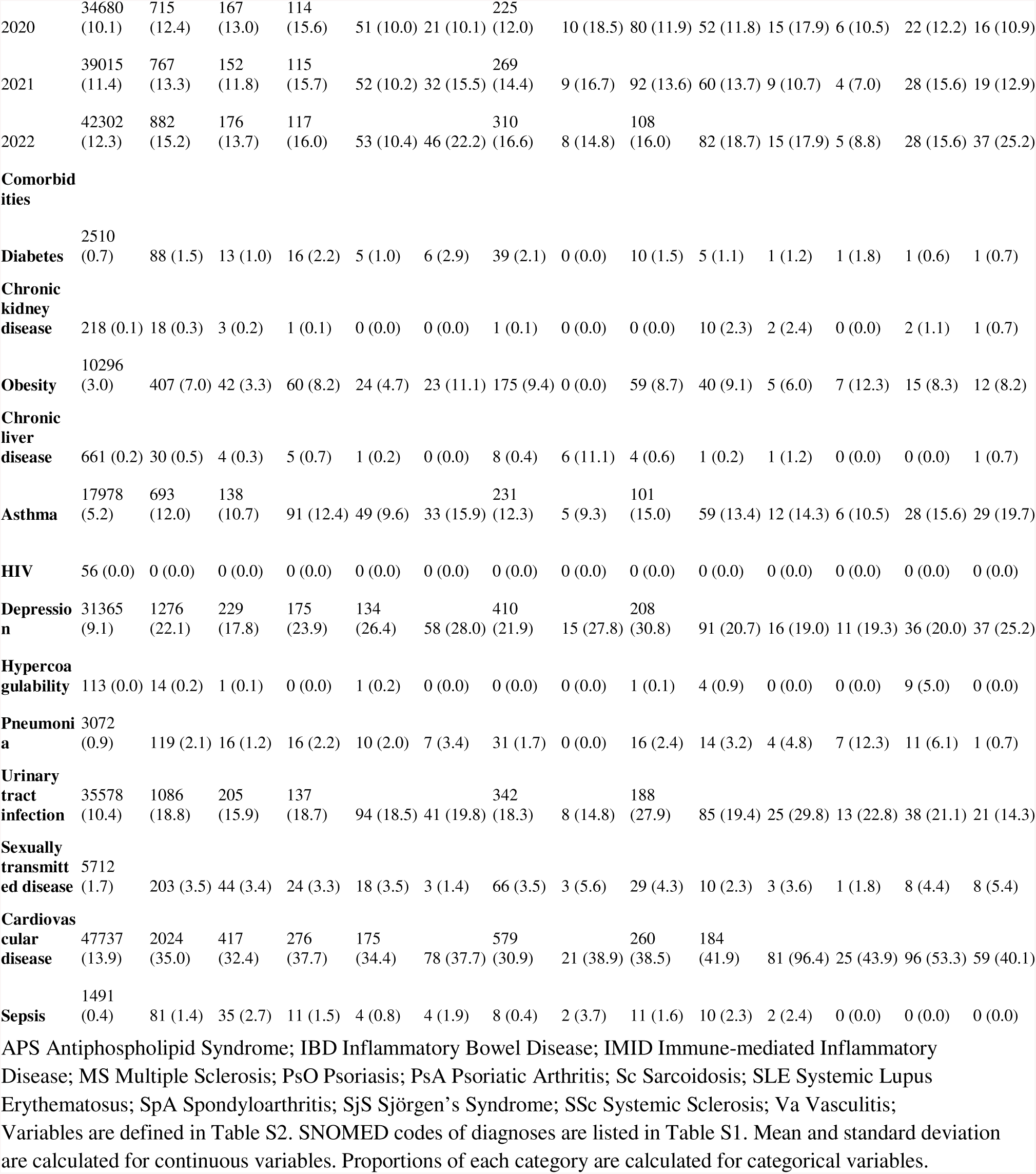
Descriptive statistics of the non-IMIDs, IMIDs, and twelve individual IMID groups.

Figure 2 describes the IMIDs cohort. 93% of people with IMIDs had one IMID diagnosis, and 7% of them had more than one IMID diagnosis (Figure 2). Figure 3 characterizes the IMMs prescription pattern of the IMIDs cohort. 83% of IMIDs people had no prenatal IMMs prescription (Figure 3A). Among the individual IMIDs we investigated, people with SLE and PsO had the highest and lowest IMMs prescription rates of 39·4% and 6·7% (Figure 3B; SLE 39·4%, RA 32·1%, SjS 31·3%, IBD 27·8%, Va 21·4%, PsA 20·8%, Sc 19·3%, APS 15·6%, SSc 14·8%, MS 12·6%, SpA 10·8%, PsO 6·7%). Among patients with IMIDs, steroids, hydroxychloroquine, 5-ASA, and TNF inhibitors were the most commonly prescribed prenatal IMMs. Prescription rates were 8%, 5%, 4%, and 3% (Figure 3C). The ranking of IMMs prescription rate varied depending on the type of disease (Figure S1). For further investigation, we focused on these four medications as the rest had a prescription rate below 2%. The prescription rate of these IMMs increased in the first trimester when compared to that of pre-pregnancy (Figure 3E). Most people prescribing these IMMs before the pregnancy continued taking them with a continuation rate ranging from 49% to 70% (Figure 3D).

**Figure 2.**
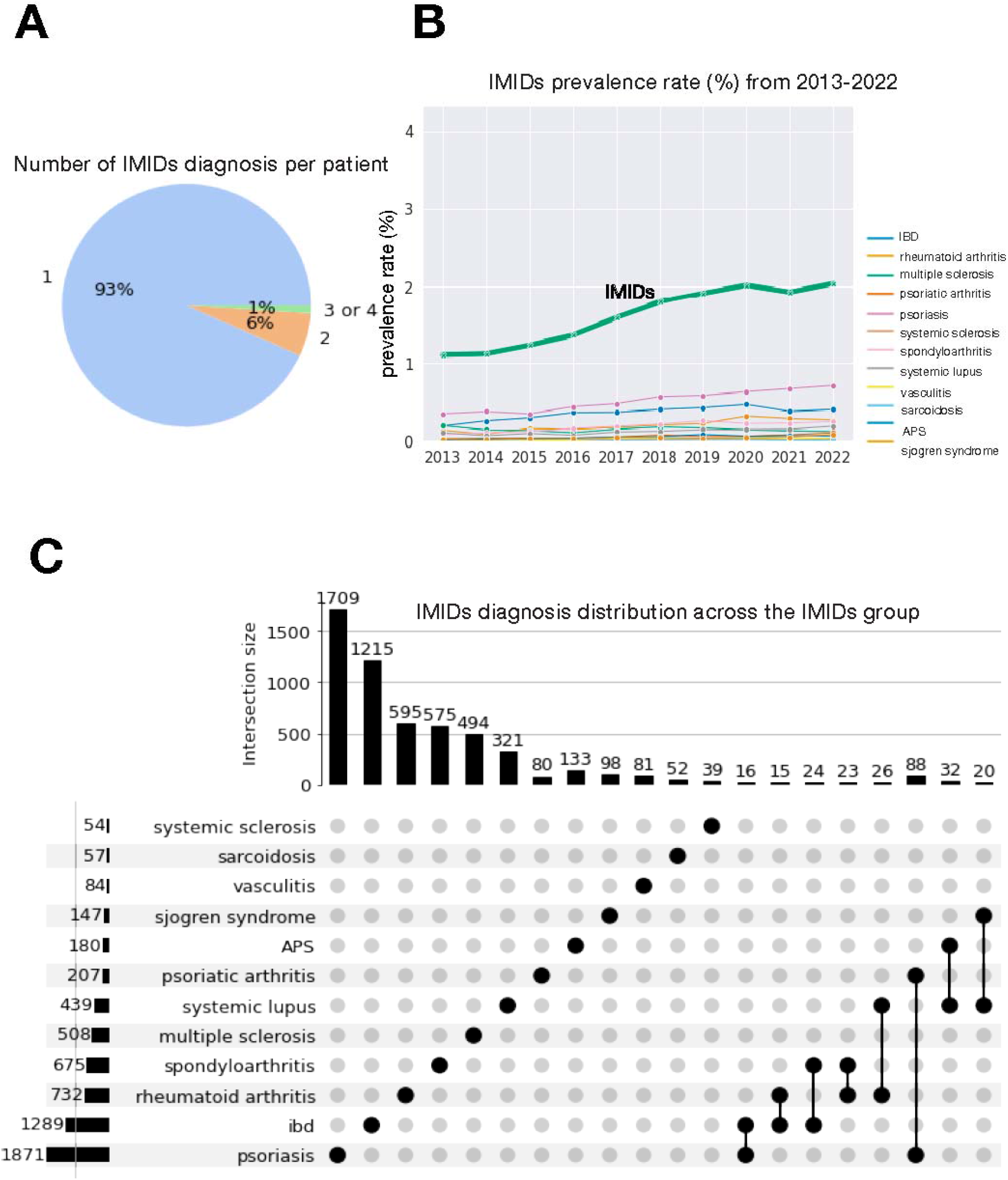
Characteristics of the IMIDs group. APS Antiphospholipid Syndrome; IBD Inflammatory Bowel Disease; IMIDs Immune-mediated Inflammatory Disease A. Number of IMIDs diagnosis per patient. 93% of the IMIDs group had one IMID diagnosis. 7% of the IMIDs group had more than one IMID diagnosis. B. IMIDs prevalence rate over time from 2013 to 2022. Prevalence rate was the proportion of IMIDs patients delivered among patients delivered in the corresponding year. IMIDs prevalence rate gradually increased from 1% to 2% from 2013 to 2022, except in the year 2021. C. IMIDs diagnosis distribution across the IMIDs group. Subsets size below 15 were not displayed. The most common and least common diagnosis was psoriasis(n=1871) and systemic sclerosis(n=54), respectively.

**Figure 3.**
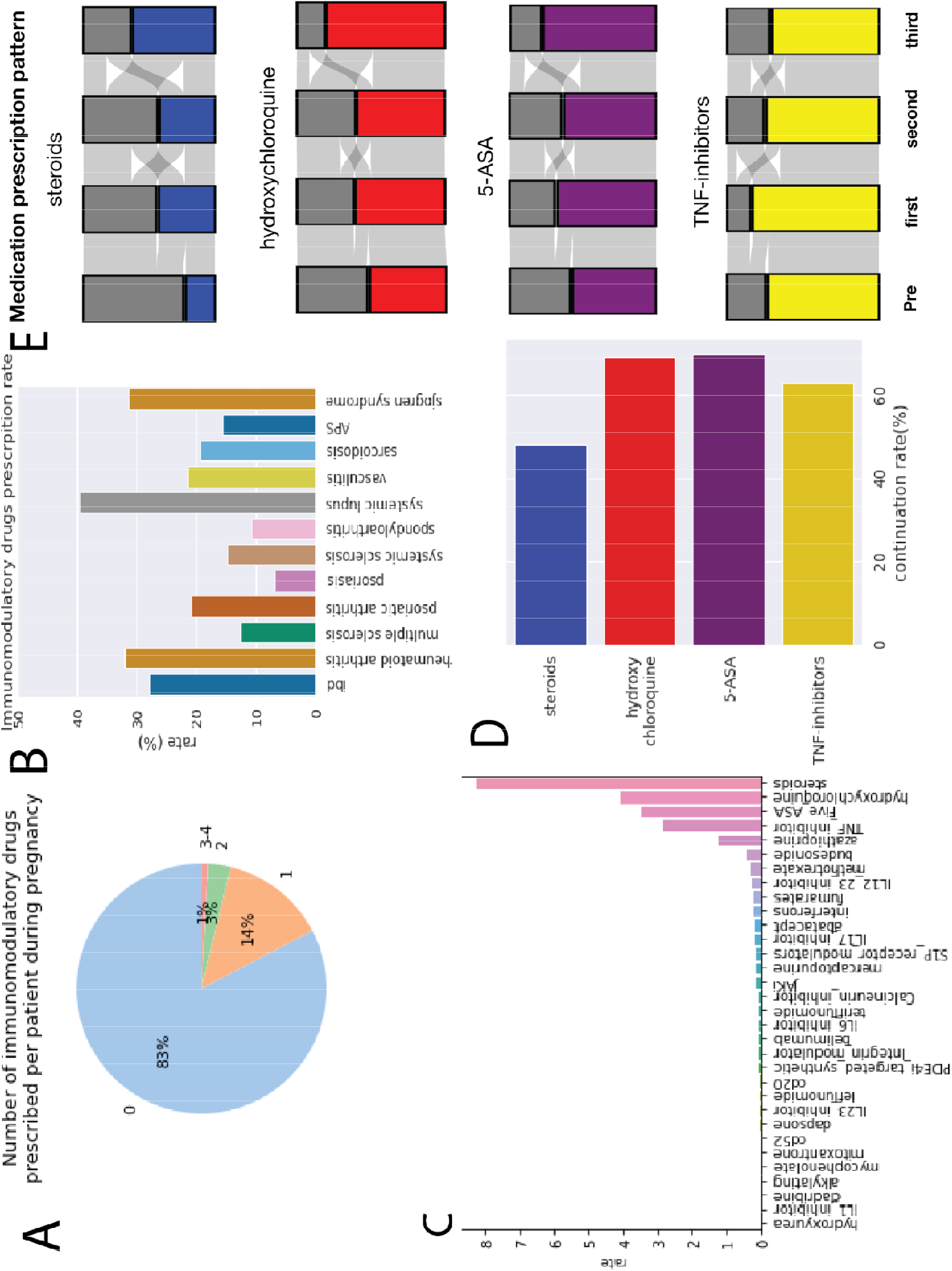
IMMs prescription pattern of the IMIDs group. APS Antiphospholipid Syndrome; IBD Inflammatory Bowel Disease; IMID Immune-mediated Inflammatory Disease; IMMs Immunomodulatory Medications; LMP Last Menstrual Period; MS Multiple Sclerosis; PsO Psoriasis; PsA Psoriatic Arthritis; Sc Sarcoidosis; SLE Systemic Lupus Erythematosus; SpA Spondyloarthritis; SjS Sjörgen’s Syndrome; SSc Systemic Sclerosis; Va Vasculitis; A. Number of IMMs prescribed per patient during pregnancy. 83% of the IMIDs group did not have any IMMs prescription. 17% had at least one IMMs prescription during pregnancy. B. Prenatal IMMs prescription rate of individual IMID groups. The descending order of prenatal IMMs prescription rate of individual IMID groups were SLE(39·4%), RA(32·1%), SjS(31·3%), IBD(27·8%), Va(21·4%), PsA(20·8%), Sc(19·3%), APS(15·6%), SSc(14·8%), MS(12·6%), SpA(10·8%), and PsO(6·7%). C. Prenatal IMMs prescription rate of the IMIDs group based on the type of IMMs. Glucocorticoids(steroids), hydroxychloroquine, 5-ASA, and TNF-α inhibitors were most commonly prescribed prenatally among the IMIDs group. Prenatal prescription rates were 8%, 5%, 4%, and 3%. Prenatal IMMs prescription rates of individual IMID groups based on the type of IMMs are displayed in Figure S1. D. IMMs continuation rate. Majority of patients, who were exposed to IMMs during 180 days prepregnancy period, continued their prescription throughout the delivery. Continuation rates ranged from 48 to 70%. E. IMMs prescription patterns among patients who prescribed corresponding IMMs at least once from LMP-180 days to delivery date. Pre, first, second, and third columns indicate 180 days prepregnancy period, first second and third trimester. Colored and gray portions respectively indicate exposed and unexposed patients for corresponding time periods.

### Association between IMIDs and risk of adverse pregnancy outcomes

When we controlled for comorbidities, having an IMID was only weakly associated with the risk of PTB (RR = 1·1[1·0, 1·3]), LBW (RR = 1·2 [1·0, 1·4]), SGA (RR = 1·1 [1·0,1·2]), and cesarean section (RR = 1·1 [1·1, 1·2]) (Figure 4). Of those 12 individual IMIDs, SpA, SLE, and APS were associated with increased risk of PTB (SpA RR = 1·5 [1·0, 2·2]; SLE RR = 2·4 [1·6, 3·6]; APS RR = 2·1 [1·2, 3·8]). SLE was the only IMID significantly correlated with enhanced risk of LBW (RR = 3·5 [2·1, 5·8]). People with RA and SLE were 1·3 ([1·0, 1·6]) and 1·9 ([1·4, 2·6]) times more likely to deliver babies with SGA condition. Patients with IBD, RA, PsA, SpA, SLE, APS, and SjShad a high likelihood chance of to deliver babies in cesarean section (IBD RR = 1·3 [1·1, 1·4], RA RR = 1·2 [1·0, 1·4], PsA RR = 1·3 [1·0, 1·8], SpA RR = 1·3 [1·1, 1·5], SLE RR = 1·3 [1·1, 1·5], APS RR = 1·7 [1·3, 2·2], SjS RR = 1·5 [1·1, 2·1]).

**Figure 4.**
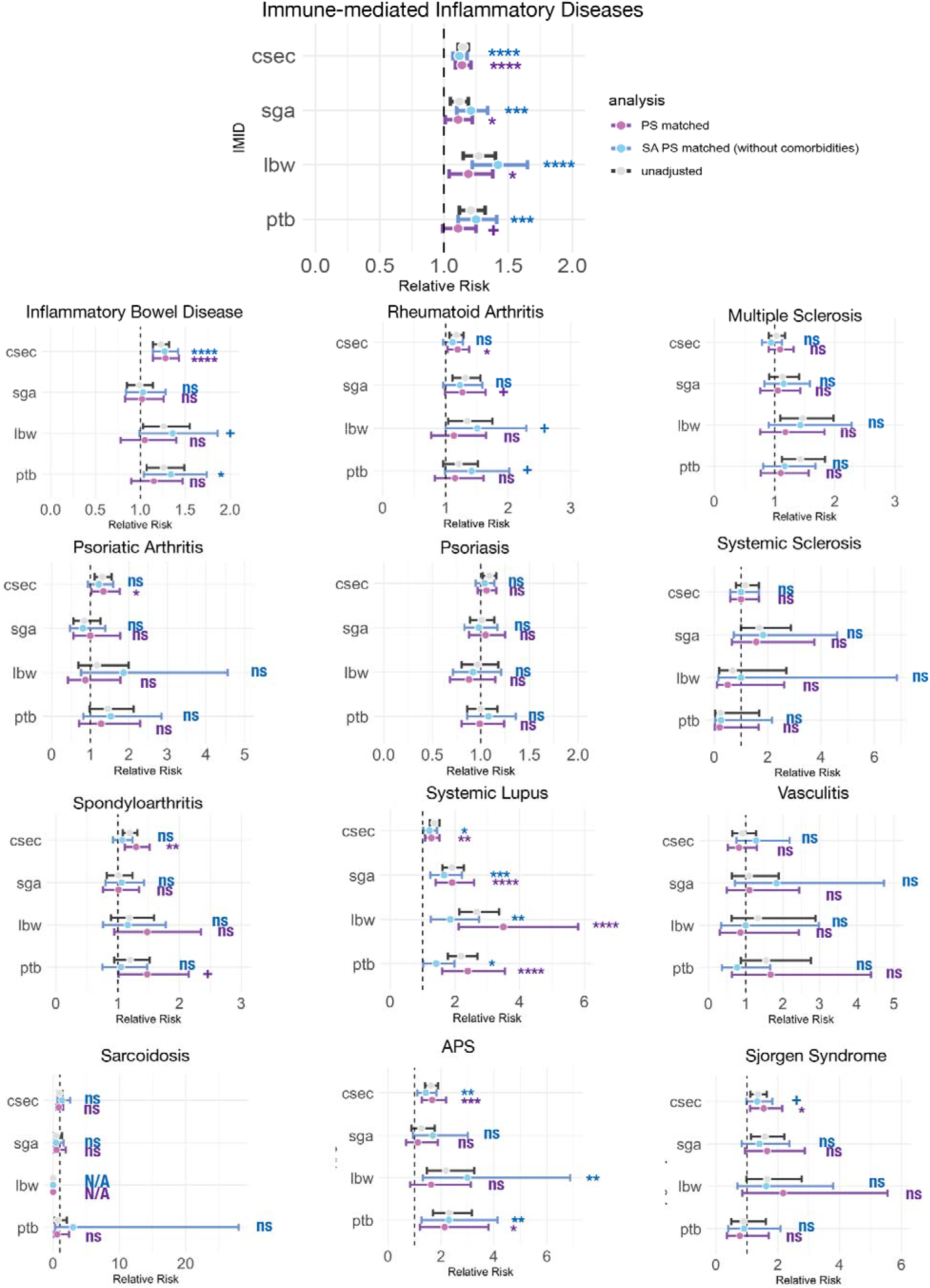
Adverse pregnancy outcomes of the IMIDs group, propensity score matched non-IMIDs group, and sensitivity analysis propensity score matched non-IMIDs group. APS Antiphospholipid Syndrome; Csec cesarean section; LBW Low Birth Weight; PTB Preterm Birth; IBD Inflammatory Bowel Disease; IMID Immune-mediated Inflammatory Disease; IMMs Immunomodulatory Medications; LMP Last Menstrual Period; MS Multiple Sclerosis; PsO Psoriasis; PsA Psoriatic Arthritis; RR Relative Risk; Sc Sarcoidosis; SGA Small for Gestational Age; SLE Systemic Lupus Erythematosus; SpA Spondyloarthritis; SjS Sjörgen’s Syndrome; SSc Systemic Sclerosis; Va Vasculitis; Sensitivity analysis was performed to assess the influence of comorbidities on the association between IMIDs and risk of adverse pregnancy outcomes. The IMIDs group had slightly elevated risk of PTB (RR=1·1[1·0,1·3]), LBW (RR=1·2[1·0,1·4]), SGA (RR=1·1[1·0,1·2]), and c-section (RR=1·1[1·1,1·2]). Of 12 individual IMID, SpA, SLE, APS was associated with increased risk of PTB (SpA RR=1.5 [1.0,2.2]; SLE RR=2.4[1.6,3.6]; APS RR=2·1[1·2,3·8]). SLE was the only IMID correlated with enhanced risk of LBW (RR=3·5[2·1,5·8]). RA and SLE patients were 1·3([1·0,1·6]) and 1.9([1·4,2·6]) times more likely to deliver SGA babies. IBD, RA, PsA, SpA, SLE, APS, and SjS patients had elevated likelihood of cesarean section delivery(IBD RR=1·3[1·1,1·4], RA RR=1·2[1·0,1·4], PsA RR=1·3[1·0,1·8], SpA RR=1·3[1·1,1·5], SLE RR=1·3[1·1,1·5], APS RR=1.7[1·3,2·2], SjS RR=1·5[1·1,2·1]). When the comorbidities were not controlled, the IMIDs group’s risk of PTB and LBW increased by 0.2. In addition, the risk of PTB and LBW of IBD and RA patients increased and gained statistical significance (IBD: PTB RR = 1·3[1·0, 1·7], LBW RR = 1·4[1·0, 1·9]; RA: PTB RR = 1·4[1·0, 2·0], LBW RR = 1·5[1·0, 2·3]). The association between APS and the risk of LBW also elevated and became statistically significant (APS LBW RR = 3·0[1·3, 6·9]). Statistical significance was reported as follows. p<0·0001:****, 0·0001≤p<0·001:***, 0·001≤p<0·01:**, 0·01≤p<0·05:*, 0·05≤p<0·1:+, 0.1<p:ns

When we did not control comorbidities to assess the impact of comorbidities on associations between IMIDs and adverse pregnancy outcomes, having any IMIDs showed 0.2 higher RR of PTB and LBW. In addition, the risk of PTB and LBW of IBD and RA patients was higher and showed statistical significance (IBD: PTB RR = 1·3[1·0, 1·7], LBW RR = 1·4[1·0, 1·9]; RA: PTB RR = 1·4[1·0, 2·0], LBW RR = 1·5[1·0, 2·3]). Association between APS and the risk of LBW also elevated and became statistically significant (APS LBW RR = 3·0[1·3, 6·9]).

## Discussion

To our knowledge, this is the first study that extensively evaluated the influence of IMIDs and comorbidities on women undergoing pregnancy. Here, we retrospectively assessed people diagnosed with one or more IMIDs before pregnancy. People who had IMIDs before pregnancy doubled from 2013 to 2022. Of the 5,784 pregnant people with IMIDs, only 17% were prescribed IMMs prenatally. Among people exposed to IMMs before the pregnancy, the majority, 48% to 70%, continued their prescriptions until delivery. People who had an IMID diagnosis before pregnancy were more likely to have comorbidities than those who did not. Overall, IMID patients had a similar but slightly elevated risk of adverse pregnancy outcomes after controlling for covariates when compared to controls. Patients with SLE had significantly elevated risk for all adverse pregnancy outcomes investigated. Out of the twelve selected IMIDs only three had an increased risk for PTB, one had an increased risk for LBW, two had an increased risk for SGA, and six had an increased risk for cesarean section. The risk of LBW and PTB of IMIDs was lower in analyses that controlled for comorbidities. We noted PTB and LBW risk on patients with RA or IBD are similar to patients without those conditions, unlike prior studies.

Prevalence rate of pregnancy with a history of IMIDs diagnosis doubled in the recent ten years. There are several possible explanations for this trend. First, maternal age increased. According to U.S. Census Bureau^22^, fertility rate of 15-29 age range gradually dropped, whereas that of the 30-44 age range rose from 1990 to 2019. As IMIDs usually develop during the reproductive age and are untreatable, older people have a higher likelihood to have IMIDs at the time of pregnancy. Second, enhanced understanding about classification and diagnosis of IMIDs could partially explain the trend. This explanation was considered as a possible reason for increase in IMIDs prevalence rate in the general population.^23^ Last but not least, it could be attributable to actual increase in the IMIDs prevalence rate. Although the exact reason of prevalence rate is not identified, there is a consensus that IMIDs prevalence is gradually increasing.^24^

We found a strong association between specific IMIDs (SLE and APS) and adverse pregnancy outcomes. This finding is consistent with previous observations that SLE and APS are risk factors for adverse pregnancy outcomes. Gao et al., 2019^12^ built a deep learning model and predicted SLE as one of the most important features in predicting extremely preterm birth. A study conducted on the largest multicentric study to prospectively assess adverse pregnancy outcomes in SLE and/or APS pregnant patients (n=385) reported nearly a 20% chance of adverse pregnancy outcomes, regardless of disease activity.^13^ Adverse pregnancy outcomes of that study were PTB, fetal/neonatal death, and fetal growth restriction. A meta-analysis of eleven observational case-control studies and thirteen cohort studies concluded that patients with SLE had twice the risk of delivering preterm when compared to controls.^14^

Surprisingly, most individual IMIDs (9: PTB, 11: LBW, 9: SGA, 6:cesarean section) were not correlated with adverse pregnancy outcomes. Given that autoimmune conditions are often considered a risk factor for high-risk pregnancy, this finding was somewhat unexpected compared to recent studies.^25, 26^ A meta-analysis conducted on twelve studies and 3907 IBD patients showed a correlation between IBD and higher incidence of PTB, LBW, and cesarean section.^16^ Huang et al., 2023^17^ evaluated eighteen studies to assess maternal outcomes of pregnant patients with RA. That study reported elevated odds of PTB, SGA, LBW, and stillbirth among patients with RA. Our study differed from these studies because we extensively controlled for comorbidities, addressing both physical and mental conditions. IBD and RA correlation with PTB and LBW decreased and lost statistical significance when we included comorbidities in the covariates used for propensity score matching. This suggeststhat comorbidities significantly influence the relationship between IBD and RA, and PTB and LBW. Our result implies that, for several IMIDs, underlying comorbidities may be more significant risk factors than IMID itself. At the same time, the low rate of medication use in this cohort may suggest that the cohort had less severe cases of IMIDs, than populations studied in academic centers. To further understand IMIDs’ relationship with adverse pregnancy outcomes, more research is needed to understand the trajectories of both IMIDs and comorbidities prior to pregnancy.

We observed a relatively low IMMs prescription rate and a high continuation rate. A meta-analysis conducted on 39 studies characterizing the use of TNF-α inhibitors among pregnant patients with IMIDs condition noted that 6% of them (1786/34223) were exposed to TNF-α inhibitors during pregnancy.^27^ This finding somewhat accords with the prenatal TNF-α inhibitors prescription rate of our findings, 2.8%, considering that the majority of their samples were IBD patients (58% vs. 22%). IMM prescription rate of patients with IBD was 1.6 times higher than for patients with IMIDs overall. The high continuation rate of IMM corresponds with that prior study. Allen et al. 2022, ^28^ a study on 338 patients with IMIDs, reported a 78% continuation rate of biologics, mostly TNF-α inhibitors. Our study found a similar rate of 70%.

The main strength of this study was the large sample size and use of longitudinal clinical observations from EHR data. We had some of the largest sample sizes of retrospective EHR records investigating the relationship between several IMIDs and pregnancy. EHR contains comprehensive and longitudinal information including but not limited to medical history, diagnosis, prescription, location of residence, and socioeconomic factors. We could control for many factors known to be associated with adverse pregnancy outcomes and IMIDs factors, including maternal and pregnancy characteristics comorbid health conditions and socioeconomic environmental factors,.

To reduce surveillance bias, we limited our analyses to continuously enrolled patients. People transiently enrolled for pregnancy are likely to lack information on medical history. Whereas, people with IMIDs may be more likely to receive more continuous care from the health system.. Therefore, people with IMIDs have more detailed information on their health condition when compared to controls, leading to surveillance bias. In addition, we further addressed this bias by conducting propensity score matching when evaluating the outcomes. The propensity score matching method matches individual people with IMID to people with similar characteristics except for the exposure to IMIDs.

A significant limitation of this study is that we did not investigate the influence of IMM s on adverse pregnancy outcomes as structured EHR cdata does not provide clear measure for disease activity. We assumed people remaining on prescription were more likely to be experiencing ongoing disease activity, affiliated with a higher chance of adverse pregnancy outcome,^29, 30^ which can induce confounding by indication. Risks and benefits of IMMs exposure could be better addressed with either natural language processing on free-text notes, or through prospective studies. Another limitation is that we considered the individual an independent entity, ignoring correlations among multiple pregnancies of a single person. As an alternative, we matched on parity and gravidity when generating matched control.

## Conclusion

Association between IMIDs and increased risk of adverse pregnancy outcome depended on the specific type of IMID and presence of comorbidities. Patients with RA or IBD had a similar likelihood to deliver preterm and low birthweight babies. There is a need to revisit the current guideline on patients with RA and IBD. We also need to include comorbidities into consideration when designing a research to investigate IMIDs on maternal health.

## Data sharing

All clinical logic has been shared. Results have been aggregated and reported within this paper to the extent possible while maintaining privacy from personal health information (PHI) as required by law. All data is archived within PSJH systems in a HIPAA-secure audited compute environment to facilitate verification of study conclusions.

## Data Availability

Results have been aggregated and reported within this paper to the extent possible while maintaining privacy from personal health information (PHI) as required by law. All data is archived within Providence Health Services and Affiliates systems in a HIPAA-secure audited compute environment to facilitate verification of study conclusions. All clinical logic for data extraction has been shared within the paper and supplemental materials. Code is publicly available at https://github.com/Hadlock-Lab/IMIDs_Maternity_Stats

https://github.com/Hadlock-Lab/IMIDs_Maternity_Stats

## Acknowledgements

We are grateful to the Institute for Systems Biology for startup funds, and to PHSA for sharing their data engineering expertise and computational resources. We would also like to acknowledge SNOMED International for developing and maintaining SNOMED-CT©.

## Author contributions

Yeon Mi Hwang: Conceptualization, Resources, Data Curation, Software, Formal Analysis, Investigation, Visualization, Data Interpretation, Methodology, Writing - original draft, Writing - review and editing; Qi Wei: Conceptualization, Data curation, Data Interpretation, Writing - original draft, Writing - review and editing; Samantha N. Piekos; Conceptualization, Data Curation, Data Interpretation, Writing - review and editing; Bhargav Vemuri: Conceptualization, Data Curation, Writing - review and editing; Sevda Molani; Conceptualization, Writing - review and editing; Philip Mease: Data Interpretation, Writing - review and editing; Leroy Hood: Conceptualization, Writing - review and editing; Jennifer J Hadlock: Conceptualization, Supervision, Writing - review and editing; YH, QW, BV, and JJH had full access to the data in the study. All authors had final responsibility for the decision to submit for publication

