## Supplementary materials for "Maternal-fetal outcomes in patients with immune mediated inflammatory diseases, with consideration of comorbidities: a retrospective cohort study in a large U.S. healthcare system"

**Supplementary Methods	1**

CDC Social Vulnerability index (CDC-SVI) and rural-urban classification	1

**Supplementary Tables	2**

Table S1. SNOMED codes for diagnoses.	2

Table S2. Variable definitions	7

Table S3. RxNorm codes for medications.	8

Table S4. IMMs prescription pattern	9

Table S5 Statistical significance of descriptive statistics	16

Table S6. Standardized mean differences after propensity score matching for individual IMIDs group	18

Table S7 Standardized mean difference after propensity score matching for individual IMIDs group in the sensitivity analysis	19

Table S8 Full list of packages used and their version number	21

**Supplementary Figures	22**

Figure S1. Prenatal IMM prescription rate of individual IMIDs	22

Figure S2. Standardized mean difference before and after propensity score matching for the IMIDs group	24

Figure S3. Hydroxychloroquine usage pattern from 2020 to 2022	24

**References	25**

### **Supplementary Methods**

#### **CDC Social Vulnerability index (CDC-SVI) and rural-urban classification**

Patient’s address was converted to a U.S. census tract. We imported publicly available CDC-SVI^1^ and rural-urban classification^2^ resources from the CDC Agency for Toxic Substance and Disease Registry and the U.S. Department of Agriculture Economic Research Service into our HIPPA-compliant workspace. Individuals’ census tract was mapped to CDC SVI and rural-urban classification code. CDC-SVI represents the percentile ranking of each census tract on 15 social factors. Social factor themes include socioeconomic status, household composition, race/ethnicity/language, and housing/transportation. CDC scores range from 0 to 1. A higher SVI score indicates a higher vulnerability of an individual’s U.S. census tract exposed for each theme. For rural-urban classification, we specifically used SecondaryRUCACode2010 (last updated in 2019) to categorize an individual’s census tract into metropolitan (<4), micropolitan (4-6), small town(7-9), and rural (10-99).

### **Supplementary Tables**

#### **Table S1. SNOMED codes for diagnoses.**

| Diagnosis | SNOMED Concept ID |
| --- | --- |
| inflammatory bowel disease | 295046003, 733157003, 721686000, 128600008, 24829000, 56689002, 71833008, 52506002, 3815005, 50440006, 1085131000119105, 426549001, 737195007, 410485009, 414156000, 966011731000119103, 61424003, 239814006, 1085901000119101, 235664007, 8161000119106, 444546002, 235714007, 1085231000119100, 201727001, 34000006, 10743231000119101, 697969008, 24526004, 1092841000119100, 1085801000119106, 15342002, 14311001, 414153008, 397173003, 414154002, 201807008, 91390005, 56287005, 239809007, 201805000, 1085851000119105, 196987008, 732966008, 13470001, 235607002, 196977009, 70622003, 1085751000119100, 78324009, 397172008, 444548001, 722850002, 7620006, 196578009, 1144956000, 359664009, 445243001, 453720571000119100, 441971007, 64766004, 442159003, 234999001, 788718000, 38106008, 969688801000119108, 78712000, 721702009, 410484008 |
| rheumatoid arthritis | 319871000119100, 319951000119101, 319061000119100, 1073741000119109, 15691841000119102, 15691121000119103, 239792003, 11055151000119108, 16050071000119108, 402433007, 193180002, 318891000119108, 1073771000119102, 201772009, 15691081000119100, 459911000124100, 402431009, 239804002, 1156774008, 193250002, 735599007, 319031000119108, 1073681000119109, 1073781000119104, 201766009, 15691881000119107, 201784006, 201780002, 1073791000119101, 410798004, 319091000119107, 201774005, 15692041000119104, 287008006, 16606721000119107, 201785007, 239803008, 82939003, 201768005, 15691321000119101, 402426007, 402428008, 410802003, 201771002, 287006005, 410801005, 1156765006, 319051000119102, 38877003, 201778008, 143441000119108, 402427003, 59165007, 318901000119107, 28880005, 15691761000119107, 427770001, 201783000, 319141000119100, 10713006, 399923009, 1073691000119107, 1073811000119102, 318941000119109, 15686321000119106, 239791005, 1073831000119107, 318921000119103, 15691241000119101, 15687841000119108, 86219005, 15691921000119100, 16606681000119101, 201769002, 201773004, 319131000119109, 410796000, 318951000119106, 318861000119101, 54867000, 239799007, 318841000119100, 15686001000119104, 52661003, 201770001, 402434001, 15687201000119107, 318871000119107, 410800006, 33719002, 318911000119105, 15685921000119102, 410502007, 15685961000119107, 201779000, 318931000119100, 1156766007, 1073711000119105, 201767000, 201777003, 7607008, 398726004, 319121000119106, 1156817001, 319071000119106, 319081000119109, 781206002, 77522006, 201775006, 318961000119108, 15673521000119101, 318851000119103, 402432002, 62131000000107, 201791009, 1073601000119101, 1156779003, 15686281000119101, 1073761000119108, 1073611000119103, 201764007, 1073721000119103, 1149218009, 16024431000119108, 15691161000119108, 410797009, 319861000119106, 287007001, 16044751000119106, 239802003, 1156777001, 318971000119102, 1073731000119100, 57160007, 429192004, 15673361000119100, 15687321000119109, 15691801000119104, 1156778006, 201776007, 239943002, 1073821000119109, 1073701000119107, 15691721000119102, 1073751000119106, 1149219001, 300961000119108, 301051000119101, 308143008, 1073801000119100, 15691961000119105, 201789001, 398640008, 400054000, 319151000119103, 319941000119103, 319041000119104, 318881000119105, 459921000124108, 410799007, 69896004, 319011000119103, 201781003, 15692001000119101, 1156819003, 319111000119104, 129563009, 201796004 |
| multiple sclerosis | 703621006, 230372003, 439567002, 724778008, 816984002, 426373005, 425500002, 445967004, 24700007, 903741000000102, 733028000, 434321000124107, 428700003, 192929006, 766246000, 192927008, 438511000, 192926004, 703622004 |
| psoriatic arthritis | 239803008, 10629311000119107, 33339001, 200956002, 239813000, 19514005, 239802003, 239804002, 410482007, 156370009, 239812005 |
| psoriasis | 238600001, 200969003, 402316001, 402314003, 238616004, 402319008, 402317005, 402326008, 402311006, 402333008, 238611009, 200966005, 402308005, 402336000, 402331005, 238605006, 402330006, 784327005, 238608008, 200965009, 200962007, 3533007, 27520001, 200967001, 111188005, 402309002, 402324006, 402318000, 238601002, 402307000, 200968006, 402320002, 28840001, 81271001, 238617008, 238603004, 200975007, 200973000, 238602009, 200972005, 238609000, 200970002, 402312004, 37042000, 238612002, 200971003, 200977004, 402315002, 200974006, 238606007, 83839005, 402313009, 402334002, 9014002, 721538000, 200963002, 402322005, 402332003, 238607003, 402323000, 402327004, 200964008, 402325007, 238613007, 784339002, 402335001, 402310007, 402321003, 402329001, 719810000, 1052325005, 238604005, 784328000, 400069004, 402337009, 25847004, 238615000 |
| systemic sclerosis | 196133001, 322461000119108, 236502006, 403519001, 298285004, 236503001, 443872005, 403517004, 402712002, 724603009, 193252005, 774080007, 403520007, 403516008, 715401008, 444133002, 201443009, 22784002, 128461001, 403518009, 1144925008, 87442008, 403514006, 402713007, 35719004, 62382002, 51156002, 89681000119101, 89155008, 299276009, 403515007, 128460000, 31848007 |
| spondyloarthritis | 239806000, 67224007, 239808004, 724606001, 1153369009, 239810002, 235066005, 236744002, 9350004, 786077003, 239805001, 201568008, 723116002, 55146009, 15630971000119102, 239815007, 784332006, 713777005, 239811003, 15972981000119101, 1074981000119100, 201736002, 9631008, 1074971000119103, 110041000119104 |
| systemic lupus erythematosus | 25380002, 54912002, 773333003, 309762007, 724767000, 19682006, 402711009, 724781003, 239888002, 397856003, 200936003, 61458006, 403487009, 80258006, 403486000, 77753005, 238652000, 201436003, 698694005, 4676006, 239886003, 196138005, 68815009, 55464009, 54072008, 239889005, 713225000, 239890001, 76521009, 36402006, 200940007, 239887007, 11013005, 73286009, 295121000119101, 22888007, 95609003, 52042003, 707301001, 295101000119105, 95644001, 403488004, 295111000119108 |
| vasculitis | 988111000000106, 239926000, 724599009, 359789008, 128971000119101, 239936008, 402662002, 402433007, 1144932004, 53485006, 232460001, 1144914004, 38675009, 11352009, 58144006, 722020006, 239946005, 28807005, 190815001, 30911005, 1144919009, 57484009, 724783000, 75053002, 191306005, 3275009, 722191003, 310701003, 21542005, 718217000, 195353004, 724600007, 239935007, 239947001, 232369001, 155441006, 239927009, 54034003, 1144805008, 235000001, 44371002, 239922003, 403443000, 57390009, 239938009, 1149218009, 723674005, 402856005, 82275008, 239939001, 239945009, 762302008, 230732009, 239925001, 239921005 |
| sarcoidosis | 724780002, 402369000, 58870009, 735433009, 187233002, 697921005, 75403004, 64757003, 233743002, 233744008, 238676008, 402371000, 238680003, 91259005, 402370004, 55941000, 234528007, 54515008, 234524009, 233771008, 707238003, 197368002, 193195000, 232368009, 402372007, 238678009, 111937006, 233769008, 111292008, 195033009, 402380000, 238679001, 9529007, 24369008, 193251003, 37061001, 192673008, 402368008, 233770009, 361197009, 402373002, 231799005, 234526006, 233767005, 72470008, 230193008, 361198004, 400127001, 31541009, 232458003, 233768000, 21787007, 238677004, 234529004, 111936002, 233772001, 234530009, 870334009, 4416007, 234531008, 425384007, 310607007, 80941006, 238674006, 17363001, 1144986007, 234527002, 238681004, 402379003 |
| antiphospholipid syndrome | 239895006, 239894005, 774084003, 239897003, 239892009, 72161000119100, 402865003, 19267009, 26843008, 609329007 |
| Sjögren's syndrome | 196137000, 239915006, 762303003, 126766000, 83901003, 78946008, 724782005, 239912009 |

| Diabetes type 1 and 2 | 395204000, 23045005, 97341000119105, 426875007, 761000119102, 359642000, 609566000, 137931000119102, 421847006, 368581000119106, 422166005, 81531005, 713706002, 16745651000119101, 16747901000119104, 721284006, 713703005, 82551000119103, 313436004, 46635009, 16747141000119104, 190389009, 769220000, 138941000119105, 368591000119109, 138911000119106, 314902007, 368101000119109, 97621000119107, 44054006, 31321000119102, 791000119109, 428007007, 421631007, 190368000, 1531000119102, 16747661000119109, 16745531000119108, 28032008, 72061000119104, 16745051000119107, 102621000119101, 781000119106, 16746261000119108, 1501000119109, 82541000119100, 1481000119100, 10660471000119109, 16746901000119101, 110181000119105, 1521000119100, 237599002, 72051000119101, 421750000, 421779007, 16746341000119103, 199230006, 427027005, 87441000119104, 422034002, 16749661000119102, 701000119103, 199229001, 609567009, 87921000119104, 16747021000119109, 712883005, 1551000119108, 16891151000119103, 9859006, 420436000, 1491000119102, 609564002, 15936501000119100, 703137001, 237627000, 97331000119101, 138921000119104, 140391000119101, 41911000119107, 10661671000119102, 140521000119107, 789567007, 16745131000119100, 313435000, 1511000119107, 10656271000119102, 110171000119107, 420756003, 87461000119100, 16745931000119102, 722454003, 789568002, 703138006, 16746131000119105, 368521000119107, 314903002, 190372001, 111231000119109, 60951000119105, 104961000119108, 16748141000119100, 28331000119107, 427134009 |
| --- | --- |
| chronic kidney disease | 731000119105, 709044004, 324311000000101, 71701000119105, 431856006, 90751000119109, 118781000119108, 127991000119101, 691421000119108, 286371000119107, 284981000119102, 104931000119100, 722150000, 285871000119106, 8501000119104, 751000119104, 324211000000106, 700378005, 90771000119100, 49708008, 950291000000103, 723190009, 368461000119103, 236436003, 897311008, 324251000000105, 285841000119104, 236435004, 10757481000119107, 324341000000100, 96441000119101, 713313000, 284971000119100, 950061000000103, 949881000000106, 714153000, 434431000124103, 46177005, 111411000119103, 949401000000103, 897312001, 722149000, 140101000119109, 90741000119107, 433146000, 324471000000100, 449631000124102, 285061000119106, 129161000119100, 153891000119101, 90761000119106, 324121000000109, 711000119100, 128001000119105, 949481000000108, 950251000000106, 285861000119100, 96741000119109, 96721000119103, 368421000119108, 708975004, 285001000119105, 722098007, 897308007, 96731000119100, 425369003, 324441000000106, 431855005, 950231000000104, 285881000119109, 950311000000102, 90731000119103, 1801000119106, 741000119101, 950181000000106, 140121000119100, 284991000119104, 96751000119106, 949921000000100, 722467000, 949421000000107, 949521000000108, 140111000119107, 140131000119102, 129181000119109, 153851000119106, 90791000119104, 285831000119108, 90721000119101, 16726004, 117681000119102, 236433006, 71421000119105, 324371000000106, 897310009, 712487000, 704667004, 285851000119102, 285041000119107, 368431000119106, 721000119107, 700379002, 950101000000101, 285911000119109, 90688005, 324281000000104, 950081000000107, 776416004, 950211000000107, 10757401000119104, 57557005, 285081000119102, 368441000119102, 285921000119102, 707324008, 236434000, 284961000119106, 714152005, 285011000119108, 324151000000104, 120261000119101, 433144002, 96711000119105, 324501000000107, 771000119108, 324411000000105, 324181000000105, 285101000119109, 368451000119100, 129151000119102, 96701000119107, 431857002, 368471000119109, 324541000000105, 949561000000100, 949621000000109, 129171000119106, 949901000000109 |
| obesity | 292464007, 72894001, 722051004, 294493008, 295509007, 238135003, 190965006, 724137002, 415530009, 248312008, 722053001, 10750551000119100, 80660001, 238134004, 717269008, 57337005, 5036006, 770680004, 702949005, 715628009, 1076701000119104, 190966007, 82793005, 248311001, 722037004, 360566006, 721231007, 297500005, 788996008, 296526005, 238136002, 414920002, 290439001, 783719006, 83911000119104, 783549006, 414917005, 717761005, 461341000124106, 414916001, 62999006, 773663004, 293481008, 444862003, 719160009, 785722006, 298464002, 63702009, 1076711000119101, 171000119107, 238133005, 414919008, 722596001, 783556000, 763350002, 719834005, 770750002, 53146006, 414438005, 270486005, 776204008, 774102003, 238132000, 1003380001, 111036000, 15750121000119108 |
| chronic liver disease | 197286002, 103611000119102, 767809001, 1010616001, 128302006, 708198006, 768006009, 45256007, 235889003, 776981000000103, 838305005, 79720007, 1155913007, 89789003, 725940006, 235896001, 21861000, 74162007, 197303009, 37688005, 699189004, 33144001, 703866000, 715401008, 19943007, 76301009, 713370005, 713181003, 197305002, 197296006, 109819003, 123605001, 123606000, 266468003, 768127002, 235895002, 38662009, 86454000, 197301006, 328383001, 425413006, 1761006, 863957008, 76783007, 314963000, 735451005, 725939009, 723829000, 838377003, 266470007, 12368000, 10295004, 735733008, 774204006, 197291001, 16070004, 9843006, 768289009, 235897005, 31155007, 536002, 450880008, 838380002, 725416005, 78208005, 768126006, 715864007, 58282009, 197293003, 57339008, 307757001, 767810006, 197284004, 419728003, 266469006, 50167007, 197299004, 41889008, 871619002, 1116000, 1155841005, 768288001, 725938001, 31712002, 123604002, 235869004, 831000119103, 713966008, 61977001, 27156006, 66870002, 420054005, 768125005, 123717006, 43904005, 60037002, 43634002, 432908002, 716203000, 266471006, 123716002, 74669004, 15999000, 197310003, 197294009, 34736002, 16098491000119109, 371139006, 271440004, 702969000, 6183001, 1092801000119102, 89580002 |
| asthma | 266361008, 426656000, 405944004, 10676431000119103, 10742121000119104, 233678006, 56968009, 2360001000004109, 125011000119100, 195949008, 1086701000000102, 34015007, 10675991000119100, 829976001, 16584951000119101, 782513000, 786836003, 370220003, 735588005, 703953004, 734905008, 10692681000119108, 1086711000000100, 418395004, 708093000, 10674711000119105, 733858005, 707445000, 401000119107, 708090002, 93432008, 72301000119103, 233683003, 424643009, 707981009, 424199006, 10675551000119104, 10674991000119104, 404808000, 407674008, 370218001, 707513007, 404806001, 901000119100, 782520007, 10675391000119101, 703954005, 63088003, 233688007, 1103911000000103, 12428000, 735589002, 1751000119100, 708094006, 389145006, 10675911000119109, 233679003, 16055311000119107, 707512002, 10676391000119108, 707447008, 425969006, 92807009, 10675751000119107, 707979007, 370219009, 18041002, 10692761000119107, 10675871000119106, 708096008, 225057002, 125021000119107, 707444001, 195967001, 866881000000101, 59786004, 442025000, 10692721000119102, 1741000119102, 19849005, 427679007, 707980005, 404804003, 423889005, 31387002, 762521001, 11641008, 370221004, 10675471000119109, 734904007, 55570000, 124991000119109, 708095007, 707511009, 57607007, 281239006, 41553006, 195977004, 409663006, 37981002, 445427006, 427295004, 103781000119103, 10675431000119106, 426979002, 99031000119107, 233687002, 427603009, 10676511000119109, 5281000124103, 707446004, 125001000119103 |
| HIV | 735521001, 713733003, 713530002, 713325002, 186707002, 713503007, 405631006, 48794007, 714083007, 713571008, 713505000, 713260006, 91948008, 1142055000, 713349004, 81000119104, 713718006, 713497004, 15928141000119107, 713278001, 186708007, 235009000, 713695001, 771119002, 713527009, 111880001, 713545009, 713731001, 713722001, 397763006, 713881001, 230598008, 1142045004, 713734009, 713339002, 186706006, 713490002, 713897006, 840442003, 713340000, 735527002, 721166000, 713491003, 713508003, 713844000, 713526000, 90681000119107, 713487008, 713510001, 10746341000119109, 838377003, 771126002, 713299003, 713275003, 713298006, 713543002, 713845004, 276665006, 713483007, 735526006, 713444005, 95892003, 713507008, 840498003, 733834006, 398329009, 40780007, 713546005, 713523008, 713445006, 79019005, 713342008, 713341001, 735525005, 421020000, 235726002, 713316008, 713489006, 735523003, 86406008, 713696000, 713967004, 713533000, 713570009, 713964006, 719522009, 722557007, 713446007, 733835007, 713572001, 713880000, 315019000, 80191000119101, 713511002, 943041000000105, 713320007, 276666007, 713730000, 735522008, 713504001, 713318009, 230180003, 713531003, 52079000, 236406007, 91947003, 713297001, 735528007, 735524009, 713729005, 713732008, 90691000119105, 713887002, 713300006, 713484001, 713532005, 62479008, 714464009, 713506004, 10755671000119100, 713544008, 240103002, 713488003, 87117006, 771127006 |
| chronic lung disease | 1010670004, 123713005, 840351007, 33325001, 233674008, 724500003, 836477007, 233615002, 233675009, 18354001, 4981000, 313299006, 47938003, 417688002, 195958001, 846637007, 789574002, 70756004, 430476004, 196049002, 313296004, 713525001, 771306007, 233762004, 847091000000104, 413839001, 314954002, 195957006, 1010334009, 713526000, 1751000119100, 723829000, 266404004, 16846004, 23958009, 68328006, 707553005, 704345008, 102361000119104, 10692761000119107, 196001008, 86680006, 240742009, 700250006, 46847001, 106001000119101, 699014000, 28791000119105, 2912004, 63841001, 18121009, 87433001, 266355005, 737180005, 707534000, 13645005, 135836000, 313297008, 36599006, 77690003, 195951007, 430969000, 196028003, 783182004, 432958009, 240747003, 26427008, 733171006, 37180002, 195959009, 1010333003, 66987001, 57686001, 266356006, 60805002, 708030004, 31898008, 293991000000106, 434301000124102, 460561000124109, 196026004, 840350008, 233677001, 16003001, 426437004, 47895001, 233713004, 285381006 |
| depression‡ | 36923009, 16238181000119101, 310495003, 83458005, 720453001, 63778009, 764711000000106, 718636001, 33736005, 426578000, 16265951000119109, 191629006, 191604000, 15193003, 76441001, 765176007, 16266991000119108, 16238221000119109, 762329003, 67711008, 40568001, 69392006, 442057004, 10835871000119104, 720451004, 247803002, 723928009, 15639000, 268621008, 832007, 29929003, 16264901000119109, 755321000000106, 19527009, 2506003, 25922000, 310497006, 16264621000119109, 30605009, 1086471000000103, 1089631000000109, 79298009, 724677009, 16265301000119106, 762345001, 191611001, 237349002, 596004, 281000119103, 430852001, 237350002, 38451003, 191676002, 18818009, 300706003, 1086681000000104, 723930006, 87842000, 191613003, 450714000, 724676000, 279225001, 764701000000109, 71336009, 16265061000119105, 84788008, 1086691000000102, 310496002, 94631000119100, 712823008, 85080004, 764691000000109, 719592004, 36474008, 231500002, 321717001, 16266831000119100, 755331000000108, 191616006, 104851000119103, 724678004, 39809009, 38694004, 274948002, 75084000, 42925002, 1153570009, 68019004, 133121000119109, 192080009, 1086661000000108, 35489007, 82218004, 60099002, 720454007, 58703003, 698957003, 1089641000000100, 838530009, 2618002, 720455008, 288751000119101, 66344007, 3109008, 36170009, 46244001, 77911002, 73867007, 191659001, 1089511000000100, 16264821000119108, 42810003, 191627008, 84760002, 10811161000119107, 10211000132109, 319768000, 231504006, 370143000, 14183003, 788120007, 762512002, 231499006, 191495003, 724690002, 78667006, 231542000, 191610000, 251000119105, 70747007, 720452006, 762321000, 28475009, 63412003, 191630001, 48589009, 231485007, 20250007, 764611000000100, 79842004, 320751009, 14070001, 726772006, 87512008, 10811121000119102, 1153575004, 33135002, 87414006, 719593009, 762336002, 1086671000000101, 33078009, 19694002, 83176005, 40379007, 192079006, 762339009, 191634005 |
| pneumonia | 206286008, 196112005, 1010670004, 80003002, 233608005, 78895009, 301004001, 724500003, 10311000175107, 10625391000119100, 441658007, 1149093006, 10625191000119102, 233728004, 408681003, 51530003, 35031000119100, 105977003, 72656004, 425464007, 41269000, 206287004, 471272001, 240741002, 10625711000119105, 40786001, 445096001, 129451001, 206284006, 430395005, 35339003, 434931000124106, 417688002, 3487004, 266350000, 191727003, 276695003, 420787001, 10625591000119108, 10301000175109, 789574002, 10625671000119106, 1092361000119109, 1010650005, 31561003, 50804000, 409664000, 60485005, 421047005, 10625471000119108, 124691000119101, 195902009, 1092951000119106, 713525001, 10624991000119103, 84753008, 71926009, 276693005, 75426006, 707552000, 422588002, 700249006, 64479007, 233607000, 233609002, 707503004, 72854003, 301003007, 312342009, 10291000175108, 277869007, 713526000, 233731003, 240635003, 10625111000119106, 10625751000119106, 233618000, 68409003, 723829000, 724499007, 22754005, 1033111000000104, 266404004, 44549008, 713084008, 772839003, 308906005, 55679008, 233730002, 438764004, 142931000119100, 10625071000119104, 707553005, 10625551000119103, 10625271000119109, 300999006, 704345008, 236302005, 102361000119104, 233617005, 195888009, 57702005, 240742009, 123589003, 700250006, 396286008, 385093006, 10625151000119107, 32204007, 58890000, 61884008, 41381004, 441590008, 1082721000119101, 75570004, 123591006, 421508002, 719218000, 240391001, 2523007, 35037009, 181007, 195900001, 195881003, 699014000, 10625511000119104, 8549006, 28791000119105, 206283000, 76090006, 763888005, 707508008, 195908008, 64880000, 233604007, 408680002, 53084003, 32286006, 233621003, 195911009, 724498004, 7063008, 195896004, 111900000, 733051000, 195886008, 39172002, 420544002, 123588006, 59475000, 64667001, 233624006, 441942006, 233625007, 426696003, 10625631000119108, 42004004, 430969000, 64917006, 415125002, 85469005, 7678002, 195909000, 10281000175105, 67525007, 128711000119106, 45556008, 240747003, 708031000, 409665004, 10625351000119105, 233610007, 83608006, 44274007, 276692000, 10625311000119109, 195878008, 301001009, 882784691000119100, 425996009, 16311000119108, 7548000, 278516003, 301000005, 421671002, 396285007, 38976008, 70036007, 10625231000119106, 123590007, 407671000, 371072008, 65141002, 46970008, 42680007, 64703005, 10625431000119105, 442094008, 195904005, 1087061000119106, 707507003, 34020007, 206289001, 426437004, 195889001, 187052004, 45312009, 233613009, 10625031000119102, 206285007, 10271000175107, 129452008, 301002002, 707449006, 870573008, 1010634002, 233623000, 314978007, 41207000, 38699009, 233614003, 713544008, 233713004, 66429007, 429271009, 233619008, 233622005, 233620002, 240387006, 123587001 |
| urinary tract infection | 197850006, 236706006, 66892003, 48278001, 301011002, 8776008, 721817004, 236620008, 197849006, 199110003, 95889002, 66993009, 371061003, 29500006, 7448003, 788962001, 301012009, 16916631000119104, 197761005, 422747000, 236721000, 240577006, 44412000, 301010001, 236702008, 51105006, 197927001, 85495007, 72225002, 111830007, 236378005, 44323002, 99631000119101, 36689008, 838353009, 431737008, 197764002, 713453003, 52123000, 4181000119109, 4009004, 1087041000119107, 129128006, 45816000, 27810000, 37133005, 307426000, 80375002, 1090711000000102, 275410008, 32268008, 33046001, 197760006, 426479005, 1723008, 1148932007, 197769007, 444834005, 1469007, 24868007, 429728004, 236602004, 707208009, 236622000, 236685000, 81359005, 197782004, 85884009, 1086071000119103, 1148870007, 236599009, 10812041000119103, 236683007, 423322005, 110041000119104, 425562008, 236600007, 236590008, 17121006, 431309003, 240578001, 700372006, 12301009, 368991000119100, 29864006, 197757004, 84619001, 88813005, 431308006, 312221005, 609491002, 369011000119102, 59530001, 123755001, 187144000, 240721007, 123754002, 1092831000119109, 199111004, 240574004, 312124009, 10751431000119104, 788961008, 236690002, 369001000119100, 197848003, 30116001, 123611003, 199206009, 91195006, 53991003, 236703003, 838301001, 373611003, 42231000009109, 370368007, 63302006, 236377000, 40635000, 236681009, 236682002, 75712001, 68566005, 54026009, 5945005, 186796004, 838302008, 307534009, 1084821000119103, 197926005, 197763008, 197768004, 23754003, 713886006, 236684001, 38176009, 179101003, 236406007, 48631008, 428091000, 199205008, 197903003, 424551004, 96171000119103, 236686004, 197606008, 1092371000119103, 1149232006, 1149233001, 236373001, 236375008, 22352007, 60867007, 236379002, 16271000119108, 236687008, 197928006, 236376009, 240579009, 197770008, 236374007, 16751003, 240576002, 5093001 |
| sexually transmitted disease‡ | 26135000, 197850006, 186846005, 85857008, 15677801000119103, 46699001, 237446005, 449773001, 237095000, 186946009, 402956009, 77939001, 1142043006, 60528006, 91554004, 194910001, 32735002, 1052319006, 1142092000, 402958005, 26039008, 237046000, 54825009, 37430004, 21523006, 240573005, 240564002, 239825002, 307423008, 81339006, 1092491000000102, 736686006, 59721007, 28198007, 240602008, 444834005, 762257007, 28867007, 266130009, 312955002, 237104007, 237096004, 235861001, 402942007, 20943002, 402950003, 186842007, 240586001, 55768006, 59233003, 444150000, 88813005, 240567009, 402126004, 194907008, 67125004, 82355002, 199164000, 11906007, 722520001, 202933002, 38523005, 266143009, 9941009, 276700005, 10754701000119100, 236682002, 237042003, 272262003, 402890001, 4082005, 402940004, 402889005, 237038001, 197348008, 240587005, 6267005, 4483005, 402952006, 35742006, 23975003, 402943002, 9241004, 4359001, 266128007, 90428001, 67391006, 15679681000119101, 230152000, 42746002, 240568004, 86028001, 86443005, 230735006, 713261005, 16217981000119107, 402951004, 312934004, 186850003, 58392004, 240577006, 44412000, 402947001, 240604009, 186878002, 240560006, 199166003, 61048000, 235863003, 427578006, 27681008, 266126006, 10759841000119105, 51960003, 1052322008, 65295003, 278480000, 235064008, 13731006, 192008, 33839006, 402949003, 54274001, 402941000, 240584003, 428230005, 423391007, 80604007, 58056005, 30168008, 402888002, 788975001, 104011000119109, 735515000, 46235002, 240554006, 28438004, 199161008, 278068003, 22386003, 109436001, 45058001, 29864006, 402894005, 302812006, 31015008, 151004, 1142095003, 59530001, 240552005, 88943008, 80770009, 274118001, 74372003, 402896007, 186833000, 789121002, 240572000, 76272004, 230563005, 449776009, 272006008, 12373006, 37754005, 2390000, 8098009, 237447001, 405635002, 240555007, 235062007, 240563008, 27648007, 301086002, 49923008, 39085002, 236687008, 410470003, 15685441000119102, 278481001, 240576002, 62861003, 193786000, 64102008, 36276008, 186868000, 240585002, 232313005, 194947001, 315826004, 186893003, 721583004, 40149008, 240582004, 72225002, 197967000, 402945009, 402895006, 186867005, 402944008, 232367004, 402955008, 53664003, 1052321001, 266136003, 15677841000119101, 1087021000119101, 186875004, 28572009, 240583009, 240558009, 1137693003, 240039005, 266133006, 9091006, 59819007, 10345003, 186931002, 186915005, 240603003, 721585006, 890115009, 35526001, 27075004, 65049003, 31999004, 235065009, 58227000, 1107004, 62207008, 94851000119107, 199165004, 15679561000119109, 240608007, 129670002, 27420004, 8555001, 68863007, 240578001, 52414005, 10754031000119105, 45377007, 20735004, 35089004, 197848003, 186847001, 240581006, 114881000119108, 197347003, 240575003, 198175009, 721587003, 66887000, 301990003, 11056091000119105, 866115006, 240557004, 83883001, 240561005, 277499006, 240569007, 59307008, 71011005, 240571007, 44743006, 1087051000119109, 439912007, 236766009, 236772009, 276877003, 240606006, 68764005, 402954007, 13310005, 1142031005, 12232008, 240579009, 237083000, 112121000119105, 233849007, 72083004, 440714005, 719755001, 10227000, 44568006, 42770003, 111807001, 60335002, 60893000, 3589003, 266125005, 31137003, 266127002, 82323002, 240565001, 5085001, 230182006, 15679481000119104, 17305005, 15677881000119106, 51928006, 406581000, 1087041000119107, 371237000, 27460003, 186854007, 237069002, 342381000119109, 11338007, 890114008, 1010615002, 838317007, 111806005, 19206003, 24868007, 827006, 61612001, 50970007, 1142099009, 16070004, 402946005, 192647003, 331151000119109, 15993391000119102, 197757004, 186861006, 240556008, 66281009, 80388004, 199163006, 186863009, 314840009, 410478005, 240574004, 240545008, 302813001, 82959004, 439913002, 35876006, 77028001, 76802005, 788998009, 240566000, 231858009, 59934002, 53529004, 197966009, 238419002, 15628003, 13095005, 186939000, 721589000, 87318008, 63751007, 50528008, 721588008, 54069001, 1087061000119106, 186903006, 19290004, 1621000119101, 1087001000119105, 198242009, 235032001, 713251003, 240562003, 275376007, 69595007, 75299005, 1052323003, 186877007 |
| cardiovascular disease | 49601007‡ |
| sepsis | 1089000, 79587009, 39246002, 2858002, 714083007, 238152004, 713866007, 447898000, 707271004, 700055009, 16060271000119109, 698819004, 700051000, 127251000119102, 449335002, 5085001, 700053002, 448421008, 715070000, 448482003, 8554002, 735638009, 1153534006, 1149494008, 762528007, 206379003, 16060311000119109, 436741000124105, 750511000000101, 448812000, 1149486001, 1125006, 199711007, 4089001, 448418006, 944071000000101, 449546000, 957431000000102, 449505005, 127091000119100, 127201000119101, 277639002, 721278008, 1092461000119104, 765106006, 1092591000119105, 206380000, 128381000119104, 276229008, 721104000, 235657005, 240389009, 447894003, 944061000000108, 276669000, 1149481006, 447897005, 735637004, 765107002, 770349000, 126811000119102, 448813005, 91302008, 448420009, 870588003, 443980004, 1036671000000106, 51831006, 700054008, 77206006, 196853004, 127181000119102, 447684006, 448422001, 277638005, 127191000119104, 127261000119100, 1092511000119101, 447843005, 1092531000119106, 10751351000119104, 32751000119100, 957421000000104, 127121000119101, 9012003, 609485004, 713865006, 240385003, 435181000124108, 127751000119108, 42939002, 789043007, 700052007, 449504009, 10001005, 448784003, 127231000119108, 1149482004, 371044001, 1092541000119102, 276741000, 733142005, 67465009, 721572008, 1076281000000108, 240426001, 46848006, 447841007, 272006008, 127311000119106, 199710008, 700050004, 200196001, 1092601000119103, 194394004, 1092471000119105, 447685007, 206378006, 11373009, 206376005, 762529004, 448417001, 186365005, 449083008, 1092501000119104, 448483008, 127171000119100, 448419003, 397843005, 447899008, 449084002, 22241000175107, 1092491000119106, 713854001, 127331000119101, 1092481000119108, 436751000124107, 1092571000119109, 449082003, 127241000119104, 24444009, 127141000119107, 16751003, 715071001, 10812001000119100 |

‡ indicate all descendant SNOMED codes are included

#### **Table S2. Variable definitions**

| **Category** | **Variable** | **Definitions** |
| --- | --- | --- |
| **Maternal/pregnancy characteristics** | Race | Race noted in medical record. Missing values were encoded as Unknown; American Indian or Alaska Native, Asian, Black or African American, Native Hawaiian or Other Pacific Islander, White or Caucasian, Multiracial, Other, Unknown |
|  | Ethnicity | Ethnicity noted in medical record. Missing values were encoded as unknown; Hispanic or Latino, Not Hispanic or Latino, Unknown |
|  | Maternal age | Maternal age at the start of pregnancy; 18 ~ 24, 25 ~ 29, 30 ~ 34, 35 ~ 40, 41 ~ 44 |
|  | Pregravid BMI | Pregravid Body Mass Index (kg/m2). Missing value was encoded as unknown; Underweight (<18.5 BMI), Normal (18.5 - 24.9 BMI), Overweight ( 25.0 - 29.9 BMI), Obese (>30.0) |
|  | Insurance status | Commercial, Medicaid, Medicare, Uninsured-Self-Pay |
|  | Smoker | Self-reported smoking status; 0,1 |
|  | Illegal drug use | Self-reported illegal drug use status; 0,1 |
|  | Preterm history | History of preterm delivery; 0,1 |
|  | Parity | Number of times a patient has delivered a fetus older than 20 weeks of gestation prior to the current pregnancy; 0, 1~5, 6 |
|  | Gravidity | Number of times a patient has been pregnant; 0, 1~5, 6 |
|  | Delivery year | Year of delivery; 2013~2022 |
| **Geographical features** | Vulnerability index of socioeconomic status | CDC SVI Socioeconomic (RPL_THEME1) theme ranking mapped to patient's U.S. Census tract; scores are 0-1. score of 0 and 1 indicate low and high level of social vulnerability regarding socioeconomic status |
|  | Vulnerability index of housing composition | CDC SVI Housing Composition & Disability (RPL_THEME2) theme ranking mapped to patient's U.S. Census tract; scores are 0-1. score of 0 and 1 indicate low and high levels of social vulnerability regarding housing composition. |
|  | Vulnerability index of minority status and language | CDC SVI Minority Status & Language (RPL_THEME3) theme ranking mapped to patient's U.S. Census tract; scores are 0-1. score of 0 and 1 indicate low and high levels of social vulnerability in terms of minority status and language. Census tract with high RPL_THEME3 score is enriched with residents of minority (non-White) and/or have low English language skills |
|  | Vulnerability index of housing type and transportation | CDC SVI Housing Type & Transportation (RPL_THEME4) theme ranking mapped to patient's U.S. Census tract; scores are 0-1. score of 0 and 1 indicate low and high levels of social vulnerability regarding housing type and transportation. Census tract with high RPL_THEME4 score is area of lower income housing and/or population dense housing options |
|  | Rural/urban categorization | U.S. Department of Agriculture Economic Research Service (USDA ERS) Rural-Urban Commuting Area (RUCA) codes; SecondaryRUCACode2010 (last updated in 2019) were mapped using patient U.S. Census Tract; Categorized as Metropolitan (> 4 score), Micropolitan (4 - 6 score), Small Town (7 - 9 score), Rural (10 - 99 score), or Unknown |
| **Maternal-fetal health outcomes** | Low birth weight | Infant birth weight ≤ 2,500g |
|  | Preterm birth | Infant gestational age (GA) at birth < 37 weeks |
|  | Small for gestational age | Infant birth weight < 10th percentiles for infants of same GA |
|  | C-section | Cesarean section |

CDC Center for Disease Control and Prevention; GA Gestational Age; SVI Social Vulnerability Index

#### **Table S3. RxNorm codes for medications.**

| **Drug class** | **Example of medications and ingredients** | **Ingredients RxNorm codes** |
| --- | --- | --- |
| hydroxychloroquine | hydroxychloroquine | 5521 |
| methotrexate | methotrexate | 6851 |
| leflunomide | leflunomide | 27169 |
| teriflunomide | teriflunomide | 1310520 |
| 5-ASA | balsalazide, sulfasalazine, mesalamine | 18747, 9524, 52582 |
| azathioprine | azathioprine | 1256 |
| mercaptopurine | mercaptopurine | 103 |
| mitoxantrone | mitoxantrone | 7005 |
| mycophenolate | mycophenolate | 265323 |
| calcineurin inhibitor | cyclosporine, sirolimus, tacrolimus | 3008, 35302, 42316 |
| TNF-α inhibitor | adalimumab, certolizumab pegol, etanercept, golimumab, infliximab | 327361, 709271, 214555, 819300, 191831 |
| fumarates | dimethyl fumarate, diroximel fumarate, monomethyl fumarate | 1373478, 2261783, 1546433 |
| interferons | interferon beta-1a, interferon beta-1b | 75917, 72257 |
| alkylating agent | chlorambucil, cyclophosphamide | 2346, 3002 |
| hydroxyurea | hydroxyurea | 5552 |
| dapsone | dapsone | 3108 |
| cladribine | cladribine | 44157 |
| IL-1 inhibitor | canakinumab, anakinra, rilonacept | 853491, 72435, 763450 |
| IL-6 inhibitor | sarilumab, tocilizumab, satralizumab | 1923319, 612865, 2391541 |
| IL-12/23 inhibitor | ustekinumab | 847083 |
| IL-17 inhibitor | ixekizumab, brodalumab, secukinumab | 1745099, 1872251, 1599788 |
| IL-23 inhibitor | guselkumab, tildrakizumab, risankizumab | 1928588, 2053436, 2166040 |
| abatacept | abatacept | 614391 |
| anti-BLyS | belimumab | 1092437 |
| S1P receptor modulator | siponimod, ponesimod, fingolimod, ozanimod | 2121085, 2532300, 1012892, 2288236 |
| JAK inhibitor | tofacitinib, upadacitinib, baricitinib | 1357536, 2196092, 2047232 |
| Integrin modulator | vedolizumab, natalizumab | 1538097, 354770 |
| PDE4 inhibitor | apremilast | 1492727 |
| anti-CD20 | rituximab, ocrelizumab, ofatumumab | 121191, 1876366, 712566 |
| anti-CD52 | alemtuzumab | 117055 |
| budesonide | budesonide | 19831 |
| systemic glucocorticoids | prednisone, dexamethasone, prednisolone, triamcinolone, methylprednisolone, hydrocortisone | 8638, 3264, 8638, 10759, 6902, 5492 |

#### **Table S4. IMMs prescription pattern**

|  | **IMIDs**  **(n=5784)** | **IBD**  **(n=1289)** | **RA**  **(n=732)** | **MS**  **(n=508)** | **PsA**  **(n=207)** | **Ps**  **(n=1871)** | **SSc**  **(n=54)** | **SpA**  **(n=675)** | **SLE**  **(n=439)** | **Va**  **(n=84)** | **Sc**  **(n=57)** | **APS**  **(n=180)** | **SjS**  **(n=147)** |
| --- | --- | --- | --- | --- | --- | --- | --- | --- | --- | --- | --- | --- | --- |
| **hydroxychloroquine** |  |  |  |  |  |  |  |  |  |  |  |  |  |
| prepregnancy | 123 (2.1) | 1 (0.1) | 53 (7.2) | 0 (0.0) | 0 (0.0) | 4 (0.2) | 3 (5.6) | 5 (0.7) | 63 (14.4) | 1 (1.2) | 1 (1.8) | 7 (3.9) | 15 (10.2) |
| prenatal | 235 (4.1) | 3 (0.2) | 93 (12.7) | 0 (0.0) | 2 (1.0) | 8 (0.4) | 6 (11.1) | 12 (1.8) | 133 (30.3) | 2 (2.4) | 1 (1.8) | 12 (6.7) | 36 (24.5) |
| first trimester | 144 (2.5) | 1 (0.1) | 63 (8.6) | 0 (0.0) | 1 (0.5) | 6 (0.3) | 3 (5.6) | 5 (0.7) | 72 (16.4) | 2 (2.4) | 1 (1.8) | 7 (3.9) | 23 (15.6) |
| second trimester | 141 (2.4) | 2 (0.2) | 52 (7.1) | 0 (0.0) | 2 (1.0) | 6 (0.3) | 4 (7.4) | 6 (0.9) | 77 (17.5) | 1 (1.2) | 1 (1.8) | 6 (3.3) | 26 (17.7) |
| third trimester | 190 (3.3) | 3 (0.2) | 70 (9.6) | 0 (0.0) | 1 (0.5) | 6 (0.3) | 4 (7.4) | 10 (1.5) | 114 (26.0) | 1 (1.2) | 1 (1.8) | 11 (6.1) | 32 (21.8) |
| **methrotrexate** |  |  |  |  |  |  |  |  |  |  |  |  |  |
| prepregnancy | 14 (0.2) | 0 (0.0) | 9 (1.2) | 0 (0.0) | 1 (0.5) | 4 (0.2) | 0 (0.0) | 2 (0.3) | 2 (0.5) | 0 (0.0) | 0 (0.0) | 0 (0.0) | 0 (0.0) |
| prenatal | 15 (0.3) | 0 (0.0) | 9 (1.2) | 0 (0.0) | 1 (0.5) | 4 (0.2) | 0 (0.0) | 2 (0.3) | 3 (0.7) | 0 (0.0) | 0 (0.0) | 0 (0.0) | 0 (0.0) |
| first trimester | 15 (0.3) | 0 (0.0) | 9 (1.2) | 0 (0.0) | 1 (0.5) | 4 (0.2) | 0 (0.0) | 2 (0.3) | 3 (0.7) | 0 (0.0) | 0 (0.0) | 0 (0.0) | 0 (0.0) |
| second trimester | 4 (0.1) | 0 (0.0) | 4 (0.5) | 0 (0.0) | 1 (0.5) | 0 (0.0) | 0 (0.0) | 1 (0.1) | 0 (0.0) | 0 (0.0) | 0 (0.0) | 0 (0.0) | 0 (0.0) |
| third trimester | 3 (0.1) | 0 (0.0) | 3 (0.4) | 0 (0.0) | 0 (0.0) | 0 (0.0) | 0 (0.0) | 0 (0.0) | 0 (0.0) | 0 (0.0) | 0 (0.0) | 0 (0.0) | 0 (0.0) |
| **leflunomide** |  |  |  |  |  |  |  |  |  |  |  |  |  |
| prepregnancy | 1 (0.0) | 0 (0.0) | 0 (0.0) | 0 (0.0) | 1 (0.5) | 1 (0.1) | 0 (0.0) | 1 (0.1) | 0 (0.0) | 0 (0.0) | 0 (0.0) | 0 (0.0) | 0 (0.0) |
| prenatal | 1 (0.0) | 0 (0.0) | 0 (0.0) | 0 (0.0) | 1 (0.5) | 1 (0.1) | 0 (0.0) | 1 (0.1) | 0 (0.0) | 0 (0.0) | 0 (0.0) | 0 (0.0) | 0 (0.0) |
| first trimester | 1 (0.0) | 0 (0.0) | 0 (0.0) | 0 (0.0) | 1 (0.5) | 1 (0.1) | 0 (0.0) | 1 (0.1) | 0 (0.0) | 0 (0.0) | 0 (0.0) | 0 (0.0) | 0 (0.0) |
| second trimester | 0 (0.0) | 0 (0.0) | 0 (0.0) | 0 (0.0) | 0 (0.0) | 0 (0.0) | 0 (0.0) | 0 (0.0) | 0 (0.0) | 0 (0.0) | 0 (0.0) | 0 (0.0) | 0 (0.0) |
| third trimester | 0 (0.0) | 0 (0.0) | 0 (0.0) | 0 (0.0) | 0 (0.0) | 0 (0.0) | 0 (0.0) | 0 (0.0) | 0 (0.0) | 0 (0.0) | 0 (0.0) | 0 (0.0) | 0 (0.0) |
| **teriflunomide** |  |  |  |  |  |  |  |  |  |  |  |  |  |
| prepregnancy | 3 (0.1) | 0 (0.0) | 0 (0.0) | 3 (0.6) | 0 (0.0) | 0 (0.0) | 0 (0.0) | 0 (0.0) | 0 (0.0) | 0 (0.0) | 0 (0.0) | 0 (0.0) | 0 (0.0) |
| prenatal | 3 (0.1) | 0 (0.0) | 0 (0.0) | 3 (0.6) | 0 (0.0) | 0 (0.0) | 0 (0.0) | 0 (0.0) | 0 (0.0) | 0 (0.0) | 0 (0.0) | 0 (0.0) | 0 (0.0) |
| first trimester | 3 (0.1) | 0 (0.0) | 0 (0.0) | 3 (0.6) | 0 (0.0) | 0 (0.0) | 0 (0.0) | 0 (0.0) | 0 (0.0) | 0 (0.0) | 0 (0.0) | 0 (0.0) | 0 (0.0) |
| second trimester | 2 (0.0) | 0 (0.0) | 0 (0.0) | 2 (0.4) | 0 (0.0) | 0 (0.0) | 0 (0.0) | 0 (0.0) | 0 (0.0) | 0 (0.0) | 0 (0.0) | 0 (0.0) | 0 (0.0) |
| third trimester | 1 (0.0) | 0 (0.0) | 0 (0.0) | 1 (0.2) | 0 (0.0) | 0 (0.0) | 0 (0.0) | 0 (0.0) | 0 (0.0) | 0 (0.0) | 0 (0.0) | 0 (0.0) | 0 (0.0) |
| **Five ASA** |  |  |  |  |  |  |  |  |  |  |  |  |  |
| prepregnancy | 116 (2.0) | 98 (7.6) | 14 (1.9) | 1 (0.2) | 1 (0.5) | 5 (0.3) | 0 (0.0) | 8 (1.2) | 1 (0.2) | 0 (0.0) | 1 (1.8) | 0 (0.0) | 1 (0.7) |
| prenatal | 200 (3.5) | 169 (13.1) | 25 (3.4) | 1 (0.2) | 4 (1.9) | 7 (0.4) | 0 (0.0) | 15 (2.2) | 4 (0.9) | 0 (0.0) | 1 (1.8) | 1 (0.6) | 1 (0.7) |
| first trimester | 137 (2.4) | 117 (9.1) | 17 (2.3) | 1 (0.2) | 2 (1.0) | 6 (0.3) | 0 (0.0) | 11 (1.6) | 1 (0.2) | 0 (0.0) | 1 (1.8) | 0 (0.0) | 1 (0.7) |
| second trimester | 128 (2.2) | 111 (8.6) | 13 (1.8) | 1 (0.2) | 2 (1.0) | 6 (0.3) | 0 (0.0) | 8 (1.2) | 2 (0.5) | 0 (0.0) | 0 (0.0) | 0 (0.0) | 0 (0.0) |
| third trimester | 157 (2.7) | 135 (10.5) | 16 (2.2) | 1 (0.2) | 2 (1.0) | 5 (0.3) | 0 (0.0) | 9 (1.3) | 4 (0.9) | 0 (0.0) | 0 (0.0) | 1 (0.6) | 0 (0.0) |
| **azathioprine** |  |  |  |  |  |  |  |  |  |  |  |  |  |
| prepregnancy | 40 (0.7) | 20 (1.6) | 10 (1.4) | 0 (0.0) | 0 (0.0) | 1 (0.1) | 0 (0.0) | 3 (0.4) | 8 (1.8) | 2 (2.4) | 0 (0.0) | 1 (0.6) | 2 (1.4) |
| prenatal | 72 (1.2) | 38 (2.9) | 14 (1.9) | 0 (0.0) | 0 (0.0) | 1 (0.1) | 1 (1.9) | 6 (0.9) | 19 (4.3) | 2 (2.4) | 0 (0.0) | 3 (1.7) | 2 (1.4) |
| first trimester | 50 (0.9) | 27 (2.1) | 12 (1.6) | 0 (0.0) | 0 (0.0) | 1 (0.1) | 0 (0.0) | 4 (0.6) | 10 (2.3) | 2 (2.4) | 0 (0.0) | 2 (1.1) | 2 (1.4) |
| second trimester | 40 (0.7) | 23 (1.8) | 6 (0.8) | 0 (0.0) | 0 (0.0) | 1 (0.1) | 0 (0.0) | 4 (0.6) | 11 (2.5) | 1 (1.2) | 0 (0.0) | 2 (1.1) | 2 (1.4) |
| third trimester | 49 (0.8) | 26 (2.0) | 7 (1.0) | 0 (0.0) | 0 (0.0) | 0 (0.0) | 1 (1.9) | 6 (0.9) | 16 (3.6) | 1 (1.2) | 0 (0.0) | 2 (1.1) | 2 (1.4) |
| **mercaptopurine** |  |  |  |  |  |  |  |  |  |  |  |  |  |
| prepregnancy | 2 (0.0) | 2 (0.2) | 0 (0.0) | 0 (0.0) | 0 (0.0) | 0 (0.0) | 0 (0.0) | 0 (0.0) | 0 (0.0) | 0 (0.0) | 0 (0.0) | 0 (0.0) | 0 (0.0) |
| prenatal | 7 (0.1) | 7 (0.5) | 0 (0.0) | 0 (0.0) | 0 (0.0) | 0 (0.0) | 0 (0.0) | 1 (0.1) | 0 (0.0) | 0 (0.0) | 0 (0.0) | 0 (0.0) | 0 (0.0) |
| first trimester | 3 (0.1) | 3 (0.2) | 0 (0.0) | 0 (0.0) | 0 (0.0) | 0 (0.0) | 0 (0.0) | 0 (0.0) | 0 (0.0) | 0 (0.0) | 0 (0.0) | 0 (0.0) | 0 (0.0) |
| second trimester | 4 (0.1) | 4 (0.3) | 0 (0.0) | 0 (0.0) | 0 (0.0) | 0 (0.0) | 0 (0.0) | 0 (0.0) | 0 (0.0) | 0 (0.0) | 0 (0.0) | 0 (0.0) | 0 (0.0) |
| third trimester | 5 (0.1) | 5 (0.4) | 0 (0.0) | 0 (0.0) | 0 (0.0) | 0 (0.0) | 0 (0.0) | 1 (0.1) | 0 (0.0) | 0 (0.0) | 0 (0.0) | 0 (0.0) | 0 (0.0) |
| **mitoxantrone** |  |  |  |  |  |  |  |  |  |  |  |  |  |
| prepregnancy | 0 (0.0) | 0 (0.0) | 0 (0.0) | 0 (0.0) | 0 (0.0) | 0 (0.0) | 0 (0.0) | 0 (0.0) | 0 (0.0) | 0 (0.0) | 0 (0.0) | 0 (0.0) | 0 (0.0) |
| prenatal | 0 (0.0) | 0 (0.0) | 0 (0.0) | 0 (0.0) | 0 (0.0) | 0 (0.0) | 0 (0.0) | 0 (0.0) | 0 (0.0) | 0 (0.0) | 0 (0.0) | 0 (0.0) | 0 (0.0) |
| first trimester | 0 (0.0) | 0 (0.0) | 0 (0.0) | 0 (0.0) | 0 (0.0) | 0 (0.0) | 0 (0.0) | 0 (0.0) | 0 (0.0) | 0 (0.0) | 0 (0.0) | 0 (0.0) | 0 (0.0) |
| second trimester | 0 (0.0) | 0 (0.0) | 0 (0.0) | 0 (0.0) | 0 (0.0) | 0 (0.0) | 0 (0.0) | 0 (0.0) | 0 (0.0) | 0 (0.0) | 0 (0.0) | 0 (0.0) | 0 (0.0) |
| third trimester | 0 (0.0) | 0 (0.0) | 0 (0.0) | 0 (0.0) | 0 (0.0) | 0 (0.0) | 0 (0.0) | 0 (0.0) | 0 (0.0) | 0 (0.0) | 0 (0.0) | 0 (0.0) | 0 (0.0) |
| **mycophenolate** |  |  |  |  |  |  |  |  |  |  |  |  |  |
| prepregnancy | 0 (0.0) | 0 (0.0) | 0 (0.0) | 0 (0.0) | 0 (0.0) | 0 (0.0) | 0 (0.0) | 0 (0.0) | 0 (0.0) | 0 (0.0) | 0 (0.0) | 0 (0.0) | 0 (0.0) |
| prenatal | 0 (0.0) | 0 (0.0) | 0 (0.0) | 0 (0.0) | 0 (0.0) | 0 (0.0) | 0 (0.0) | 0 (0.0) | 0 (0.0) | 0 (0.0) | 0 (0.0) | 0 (0.0) | 0 (0.0) |
| first trimester | 0 (0.0) | 0 (0.0) | 0 (0.0) | 0 (0.0) | 0 (0.0) | 0 (0.0) | 0 (0.0) | 0 (0.0) | 0 (0.0) | 0 (0.0) | 0 (0.0) | 0 (0.0) | 0 (0.0) |
| second trimester | 0 (0.0) | 0 (0.0) | 0 (0.0) | 0 (0.0) | 0 (0.0) | 0 (0.0) | 0 (0.0) | 0 (0.0) | 0 (0.0) | 0 (0.0) | 0 (0.0) | 0 (0.0) | 0 (0.0) |
| third trimester | 0 (0.0) | 0 (0.0) | 0 (0.0) | 0 (0.0) | 0 (0.0) | 0 (0.0) | 0 (0.0) | 0 (0.0) | 0 (0.0) | 0 (0.0) | 0 (0.0) | 0 (0.0) | 0 (0.0) |
| **calcineurin inhibitor** |  |  |  |  |  |  |  |  |  |  |  |  |  |
| prepregnancy | 1 (0.0) | 1 (0.1) | 0 (0.0) | 0 (0.0) | 0 (0.0) | 0 (0.0) | 0 (0.0) | 0 (0.0) | 0 (0.0) | 0 (0.0) | 0 (0.0) | 0 (0.0) | 0 (0.0) |
| prenatal | 4 (0.1) | 1 (0.1) | 0 (0.0) | 0 (0.0) | 0 (0.0) | 0 (0.0) | 0 (0.0) | 0 (0.0) | 3 (0.7) | 1 (1.2) | 0 (0.0) | 0 (0.0) | 1 (0.7) |
| first trimester | 1 (0.0) | 1 (0.1) | 0 (0.0) | 0 (0.0) | 0 (0.0) | 0 (0.0) | 0 (0.0) | 0 (0.0) | 0 (0.0) | 0 (0.0) | 0 (0.0) | 0 (0.0) | 0 (0.0) |
| second trimester | 1 (0.0) | 0 (0.0) | 0 (0.0) | 0 (0.0) | 0 (0.0) | 0 (0.0) | 0 (0.0) | 0 (0.0) | 1 (0.2) | 0 (0.0) | 0 (0.0) | 0 (0.0) | 0 (0.0) |
| third trimester | 2 (0.0) | 0 (0.0) | 0 (0.0) | 0 (0.0) | 0 (0.0) | 0 (0.0) | 0 (0.0) | 0 (0.0) | 2 (0.5) | 1 (1.2) | 0 (0.0) | 0 (0.0) | 1 (0.7) |
| **TNF-α inhibitor** |  |  |  |  |  |  |  |  |  |  |  |  |  |
| prepregnancy | 121 (2.1) | 38 (2.9) | 61 (8.3) | 0 (0.0) | 19 (9.2) | 22 (1.2) | 0 (0.0) | 16 (2.4) | 3 (0.7) | 1 (1.2) | 1 (1.8) | 0 (0.0) | 0 (0.0) |
| prenatal | 163 (2.8) | 52 (4.0) | 82 (11.2) | 0 (0.0) | 21 (10.1) | 23 (1.2) | 0 (0.0) | 26 (3.9) | 5 (1.1) | 1 (1.2) | 1 (1.8) | 0 (0.0) | 1 (0.7) |
| first trimester | 138 (2.4) | 45 (3.5) | 68 (9.3) | 0 (0.0) | 20 (9.7) | 23 (1.2) | 0 (0.0) | 20 (3.0) | 3 (0.7) | 1 (1.2) | 1 (1.8) | 0 (0.0) | 0 (0.0) |
| second trimester | 121 (2.1) | 41 (3.2) | 57 (7.8) | 0 (0.0) | 16 (7.7) | 20 (1.1) | 0 (0.0) | 20 (3.0) | 3 (0.7) | 0 (0.0) | 0 (0.0) | 0 (0.0) | 1 (0.7) |
| third trimester | 117 (2.0) | 44 (3.4) | 53 (7.2) | 0 (0.0) | 13 (6.3) | 18 (1.0) | 0 (0.0) | 20 (3.0) | 4 (0.9) | 0 (0.0) | 0 (0.0) | 0 (0.0) | 1 (0.7) |
| **fumarates** |  |  |  |  |  |  |  |  |  |  |  |  |  |
| prepregnancy | 9 (0.2) | 0 (0.0) | 0 (0.0) | 9 (1.8) | 0 (0.0) | 0 (0.0) | 0 (0.0) | 0 (0.0) | 0 (0.0) | 0 (0.0) | 0 (0.0) | 0 (0.0) | 0 (0.0) |
| prenatal | 11 (0.2) | 0 (0.0) | 0 (0.0) | 11 (2.2) | 0 (0.0) | 0 (0.0) | 0 (0.0) | 0 (0.0) | 0 (0.0) | 0 (0.0) | 0 (0.0) | 0 (0.0) | 0 (0.0) |
| first trimester | 9 (0.2) | 0 (0.0) | 0 (0.0) | 9 (1.8) | 0 (0.0) | 0 (0.0) | 0 (0.0) | 0 (0.0) | 0 (0.0) | 0 (0.0) | 0 (0.0) | 0 (0.0) | 0 (0.0) |
| second trimester | 5 (0.1) | 0 (0.0) | 0 (0.0) | 5 (1.0) | 0 (0.0) | 0 (0.0) | 0 (0.0) | 0 (0.0) | 0 (0.0) | 0 (0.0) | 0 (0.0) | 0 (0.0) | 0 (0.0) |
| third trimester | 5 (0.1) | 0 (0.0) | 0 (0.0) | 5 (1.0) | 0 (0.0) | 0 (0.0) | 0 (0.0) | 0 (0.0) | 0 (0.0) | 0 (0.0) | 0 (0.0) | 0 (0.0) | 0 (0.0) |
| **interferons** |  |  |  |  |  |  |  |  |  |  |  |  |  |
| prepregnancy | 9 (0.2) | 0 (0.0) | 0 (0.0) | 9 (1.8) | 0 (0.0) | 0 (0.0) | 0 (0.0) | 0 (0.0) | 0 (0.0) | 0 (0.0) | 0 (0.0) | 0 (0.0) | 0 (0.0) |
| prenatal | 10 (0.2) | 0 (0.0) | 0 (0.0) | 10 (2.0) | 0 (0.0) | 0 (0.0) | 0 (0.0) | 0 (0.0) | 0 (0.0) | 0 (0.0) | 0 (0.0) | 0 (0.0) | 0 (0.0) |
| first trimester | 10 (0.2) | 0 (0.0) | 0 (0.0) | 10 (2.0) | 0 (0.0) | 0 (0.0) | 0 (0.0) | 0 (0.0) | 0 (0.0) | 0 (0.0) | 0 (0.0) | 0 (0.0) | 0 (0.0) |
| second trimester | 7 (0.1) | 0 (0.0) | 0 (0.0) | 7 (1.4) | 0 (0.0) | 0 (0.0) | 0 (0.0) | 0 (0.0) | 0 (0.0) | 0 (0.0) | 0 (0.0) | 0 (0.0) | 0 (0.0) |
| third trimester | 5 (0.1) | 0 (0.0) | 0 (0.0) | 5 (1.0) | 0 (0.0) | 0 (0.0) | 0 (0.0) | 0 (0.0) | 0 (0.0) | 0 (0.0) | 0 (0.0) | 0 (0.0) | 0 (0.0) |
| **alkylating agent** |  |  |  |  |  |  |  |  |  |  |  |  |  |
| prepregnancy | 0 (0.0) | 0 (0.0) | 0 (0.0) | 0 (0.0) | 0 (0.0) | 0 (0.0) | 0 (0.0) | 0 (0.0) | 0 (0.0) | 0 (0.0) | 0 (0.0) | 0 (0.0) | 0 (0.0) |
| prenatal | 0 (0.0) | 0 (0.0) | 0 (0.0) | 0 (0.0) | 0 (0.0) | 0 (0.0) | 0 (0.0) | 0 (0.0) | 0 (0.0) | 0 (0.0) | 0 (0.0) | 0 (0.0) | 0 (0.0) |
| first trimester | 0 (0.0) | 0 (0.0) | 0 (0.0) | 0 (0.0) | 0 (0.0) | 0 (0.0) | 0 (0.0) | 0 (0.0) | 0 (0.0) | 0 (0.0) | 0 (0.0) | 0 (0.0) | 0 (0.0) |
| second trimester | 0 (0.0) | 0 (0.0) | 0 (0.0) | 0 (0.0) | 0 (0.0) | 0 (0.0) | 0 (0.0) | 0 (0.0) | 0 (0.0) | 0 (0.0) | 0 (0.0) | 0 (0.0) | 0 (0.0) |
| third trimester | 0 (0.0) | 0 (0.0) | 0 (0.0) | 0 (0.0) | 0 (0.0) | 0 (0.0) | 0 (0.0) | 0 (0.0) | 0 (0.0) | 0 (0.0) | 0 (0.0) | 0 (0.0) | 0 (0.0) |
| **hydroxyurea** |  |  |  |  |  |  |  |  |  |  |  |  |  |
| prepregnancy | 0 (0.0) | 0 (0.0) | 0 (0.0) | 0 (0.0) | 0 (0.0) | 0 (0.0) | 0 (0.0) | 0 (0.0) | 0 (0.0) | 0 (0.0) | 0 (0.0) | 0 (0.0) | 0 (0.0) |
| prenatal | 0 (0.0) | 0 (0.0) | 0 (0.0) | 0 (0.0) | 0 (0.0) | 0 (0.0) | 0 (0.0) | 0 (0.0) | 0 (0.0) | 0 (0.0) | 0 (0.0) | 0 (0.0) | 0 (0.0) |
| first trimester | 0 (0.0) | 0 (0.0) | 0 (0.0) | 0 (0.0) | 0 (0.0) | 0 (0.0) | 0 (0.0) | 0 (0.0) | 0 (0.0) | 0 (0.0) | 0 (0.0) | 0 (0.0) | 0 (0.0) |
| second trimester | 0 (0.0) | 0 (0.0) | 0 (0.0) | 0 (0.0) | 0 (0.0) | 0 (0.0) | 0 (0.0) | 0 (0.0) | 0 (0.0) | 0 (0.0) | 0 (0.0) | 0 (0.0) | 0 (0.0) |
| third trimester | 0 (0.0) | 0 (0.0) | 0 (0.0) | 0 (0.0) | 0 (0.0) | 0 (0.0) | 0 (0.0) | 0 (0.0) | 0 (0.0) | 0 (0.0) | 0 (0.0) | 0 (0.0) | 0 (0.0) |
| **dapsone** |  |  |  |  |  |  |  |  |  |  |  |  |  |
| prepregnancy | 1 (0.0) | 0 (0.0) | 1 (0.1) | 0 (0.0) | 0 (0.0) | 0 (0.0) | 0 (0.0) | 0 (0.0) | 0 (0.0) | 0 (0.0) | 0 (0.0) | 0 (0.0) | 0 (0.0) |
| prenatal | 1 (0.0) | 0 (0.0) | 1 (0.1) | 0 (0.0) | 0 (0.0) | 0 (0.0) | 0 (0.0) | 0 (0.0) | 0 (0.0) | 0 (0.0) | 0 (0.0) | 0 (0.0) | 0 (0.0) |
| first trimester | 1 (0.0) | 0 (0.0) | 1 (0.1) | 0 (0.0) | 0 (0.0) | 0 (0.0) | 0 (0.0) | 0 (0.0) | 0 (0.0) | 0 (0.0) | 0 (0.0) | 0 (0.0) | 0 (0.0) |
| second trimester | 0 (0.0) | 0 (0.0) | 0 (0.0) | 0 (0.0) | 0 (0.0) | 0 (0.0) | 0 (0.0) | 0 (0.0) | 0 (0.0) | 0 (0.0) | 0 (0.0) | 0 (0.0) | 0 (0.0) |
| third trimester | 0 (0.0) | 0 (0.0) | 0 (0.0) | 0 (0.0) | 0 (0.0) | 0 (0.0) | 0 (0.0) | 0 (0.0) | 0 (0.0) | 0 (0.0) | 0 (0.0) | 0 (0.0) | 0 (0.0) |
| **cladribine** | 0 (0.0) | 0 (0.0) | 0 (0.0) | 0 (0.0) | 0 (0.0) | 0 (0.0) | 0 (0.0) | 0 (0.0) | 0 (0.0) | 0 (0.0) | 0 (0.0) | 0 (0.0) | 0 (0.0) |
| prepregnancy | 0 (0.0) | 0 (0.0) | 0 (0.0) | 0 (0.0) | 0 (0.0) | 0 (0.0) | 0 (0.0) | 0 (0.0) | 0 (0.0) | 0 (0.0) | 0 (0.0) | 0 (0.0) | 0 (0.0) |
| prenatal | 0 (0.0) | 0 (0.0) | 0 (0.0) | 0 (0.0) | 0 (0.0) | 0 (0.0) | 0 (0.0) | 0 (0.0) | 0 (0.0) | 0 (0.0) | 0 (0.0) | 0 (0.0) | 0 (0.0) |
| first trimester | 0 (0.0) | 0 (0.0) | 0 (0.0) | 0 (0.0) | 0 (0.0) | 0 (0.0) | 0 (0.0) | 0 (0.0) | 0 (0.0) | 0 (0.0) | 0 (0.0) | 0 (0.0) | 0 (0.0) |
| second trimester | 0 (0.0) | 0 (0.0) | 0 (0.0) | 0 (0.0) | 0 (0.0) | 0 (0.0) | 0 (0.0) | 0 (0.0) | 0 (0.0) | 0 (0.0) | 0 (0.0) | 0 (0.0) | 0 (0.0) |
| third trimester | 0 (0.0) | 0 (0.0) | 0 (0.0) | 0 (0.0) | 0 (0.0) | 0 (0.0) | 0 (0.0) | 0 (0.0) | 0 (0.0) | 0 (0.0) | 0 (0.0) | 0 (0.0) | 0 (0.0) |
| **IL1 inhibitor** |  |  |  |  |  |  |  |  |  |  |  |  |  |
| prepregnancy | 0 (0.0) | 0 (0.0) | 0 (0.0) | 0 (0.0) | 0 (0.0) | 0 (0.0) | 0 (0.0) | 0 (0.0) | 0 (0.0) | 0 (0.0) | 0 (0.0) | 0 (0.0) | 0 (0.0) |
| prenatal | 0 (0.0) | 0 (0.0) | 0 (0.0) | 0 (0.0) | 0 (0.0) | 0 (0.0) | 0 (0.0) | 0 (0.0) | 0 (0.0) | 0 (0.0) | 0 (0.0) | 0 (0.0) | 0 (0.0) |
| first trimester | 0 (0.0) | 0 (0.0) | 0 (0.0) | 0 (0.0) | 0 (0.0) | 0 (0.0) | 0 (0.0) | 0 (0.0) | 0 (0.0) | 0 (0.0) | 0 (0.0) | 0 (0.0) | 0 (0.0) |
| second trimester | 0 (0.0) | 0 (0.0) | 0 (0.0) | 0 (0.0) | 0 (0.0) | 0 (0.0) | 0 (0.0) | 0 (0.0) | 0 (0.0) | 0 (0.0) | 0 (0.0) | 0 (0.0) | 0 (0.0) |
| third trimester | 0 (0.0) | 0 (0.0) | 0 (0.0) | 0 (0.0) | 0 (0.0) | 0 (0.0) | 0 (0.0) | 0 (0.0) | 0 (0.0) | 0 (0.0) | 0 (0.0) | 0 (0.0) | 0 (0.0) |
| **IL6 inhibitor** |  |  |  |  |  |  |  |  |  |  |  |  |  |
| prepregnancy | 2 (0.0) | 0 (0.0) | 2 (0.3) | 0 (0.0) | 0 (0.0) | 0 (0.0) | 0 (0.0) | 0 (0.0) | 0 (0.0) | 0 (0.0) | 0 (0.0) | 0 (0.0) | 0 (0.0) |
| prenatal | 3 (0.1) | 0 (0.0) | 2 (0.3) | 1 (0.2) | 0 (0.0) | 0 (0.0) | 0 (0.0) | 0 (0.0) | 0 (0.0) | 0 (0.0) | 0 (0.0) | 0 (0.0) | 0 (0.0) |
| first trimester | 3 (0.1) | 0 (0.0) | 2 (0.3) | 1 (0.2) | 0 (0.0) | 0 (0.0) | 0 (0.0) | 0 (0.0) | 0 (0.0) | 0 (0.0) | 0 (0.0) | 0 (0.0) | 0 (0.0) |
| second trimester | 1 (0.0) | 0 (0.0) | 0 (0.0) | 1 (0.2) | 0 (0.0) | 0 (0.0) | 0 (0.0) | 0 (0.0) | 0 (0.0) | 0 (0.0) | 0 (0.0) | 0 (0.0) | 0 (0.0) |
| third trimester | 1 (0.0) | 0 (0.0) | 0 (0.0) | 1 (0.2) | 0 (0.0) | 0 (0.0) | 0 (0.0) | 0 (0.0) | 0 (0.0) | 0 (0.0) | 0 (0.0) | 0 (0.0) | 0 (0.0) |
| **IL12/IL23 inhibitor** |  |  |  |  |  |  |  |  |  |  |  |  |  |
| prepregnancy | 11 (0.2) | 8 (0.6) | 0 (0.0) | 0 (0.0) | 1 (0.5) | 3 (0.2) | 0 (0.0) | 1 (0.1) | 0 (0.0) | 0 (0.0) | 0 (0.0) | 0 (0.0) | 0 (0.0) |
| prenatal | 14 (0.2) | 10 (0.8) | 0 (0.0) | 0 (0.0) | 1 (0.5) | 4 (0.2) | 0 (0.0) | 1 (0.1) | 0 (0.0) | 0 (0.0) | 0 (0.0) | 0 (0.0) | 0 (0.0) |
| first trimester | 12 (0.2) | 8 (0.6) | 0 (0.0) | 0 (0.0) | 1 (0.5) | 4 (0.2) | 0 (0.0) | 1 (0.1) | 0 (0.0) | 0 (0.0) | 0 (0.0) | 0 (0.0) | 0 (0.0) |
| second trimester | 12 (0.2) | 10 (0.8) | 0 (0.0) | 0 (0.0) | 1 (0.5) | 2 (0.1) | 0 (0.0) | 1 (0.1) | 0 (0.0) | 0 (0.0) | 0 (0.0) | 0 (0.0) | 0 (0.0) |
| third trimester | 12 (0.2) | 10 (0.8) | 0 (0.0) | 0 (0.0) | 1 (0.5) | 2 (0.1) | 0 (0.0) | 1 (0.1) | 0 (0.0) | 0 (0.0) | 0 (0.0) | 0 (0.0) | 0 (0.0) |
| **IL17 inhibitor** |  |  |  |  |  |  |  |  |  |  |  |  |  |
| prepregnancy | 8 (0.1) | 0 (0.0) | 1 (0.1) | 0 (0.0) | 2 (1.0) | 6 (0.3) | 0 (0.0) | 2 (0.3) | 0 (0.0) | 0 (0.0) | 0 (0.0) | 0 (0.0) | 0 (0.0) |
| prenatal | 8 (0.1) | 0 (0.0) | 1 (0.1) | 0 (0.0) | 2 (1.0) | 6 (0.3) | 0 (0.0) | 2 (0.3) | 0 (0.0) | 0 (0.0) | 0 (0.0) | 0 (0.0) | 0 (0.0) |
| first trimester | 8 (0.1) | 0 (0.0) | 1 (0.1) | 0 (0.0) | 2 (1.0) | 6 (0.3) | 0 (0.0) | 2 (0.3) | 0 (0.0) | 0 (0.0) | 0 (0.0) | 0 (0.0) | 0 (0.0) |
| second trimester | 5 (0.1) | 0 (0.0) | 1 (0.1) | 0 (0.0) | 1 (0.5) | 3 (0.2) | 0 (0.0) | 2 (0.3) | 0 (0.0) | 0 (0.0) | 0 (0.0) | 0 (0.0) | 0 (0.0) |
| third trimester | 2 (0.0) | 0 (0.0) | 1 (0.1) | 0 (0.0) | 1 (0.5) | 2 (0.1) | 0 (0.0) | 0 (0.0) | 0 (0.0) | 0 (0.0) | 0 (0.0) | 0 (0.0) | 0 (0.0) |
| **IL23 inhibitor** |  |  |  |  |  |  |  |  |  |  |  |  |  |
| prepregnancy | 1 (0.0) | 0 (0.0) | 0 (0.0) | 0 (0.0) | 0 (0.0) | 1 (0.1) | 0 (0.0) | 0 (0.0) | 0 (0.0) | 0 (0.0) | 0 (0.0) | 0 (0.0) | 0 (0.0) |
| prenatal | 1 (0.0) | 0 (0.0) | 0 (0.0) | 0 (0.0) | 0 (0.0) | 1 (0.1) | 0 (0.0) | 0 (0.0) | 0 (0.0) | 0 (0.0) | 0 (0.0) | 0 (0.0) | 0 (0.0) |
| first trimester | 1 (0.0) | 0 (0.0) | 0 (0.0) | 0 (0.0) | 0 (0.0) | 1 (0.1) | 0 (0.0) | 0 (0.0) | 0 (0.0) | 0 (0.0) | 0 (0.0) | 0 (0.0) | 0 (0.0) |
| second trimester | 1 (0.0) | 0 (0.0) | 0 (0.0) | 0 (0.0) | 0 (0.0) | 1 (0.1) | 0 (0.0) | 0 (0.0) | 0 (0.0) | 0 (0.0) | 0 (0.0) | 0 (0.0) | 0 (0.0) |
| third trimester | 1 (0.0) | 0 (0.0) | 0 (0.0) | 0 (0.0) | 0 (0.0) | 1 (0.1) | 0 (0.0) | 0 (0.0) | 0 (0.0) | 0 (0.0) | 0 (0.0) | 0 (0.0) | 0 (0.0) |
| **abatacept** |  |  |  |  |  |  |  |  |  |  |  |  |  |
| prepregnancy | 8 (0.1) | 0 (0.0) | 8 (1.1) | 0 (0.0) | 0 (0.0) | 0 (0.0) | 0 (0.0) | 2 (0.3) | 2 (0.5) | 0 (0.0) | 0 (0.0) | 0 (0.0) | 0 (0.0) |
| prenatal | 9 (0.2) | 0 (0.0) | 9 (1.2) | 0 (0.0) | 0 (0.0) | 0 (0.0) | 0 (0.0) | 2 (0.3) | 2 (0.5) | 0 (0.0) | 0 (0.0) | 0 (0.0) | 0 (0.0) |
| first trimester | 9 (0.2) | 0 (0.0) | 9 (1.2) | 0 (0.0) | 0 (0.0) | 0 (0.0) | 0 (0.0) | 2 (0.3) | 2 (0.5) | 0 (0.0) | 0 (0.0) | 0 (0.0) | 0 (0.0) |
| second trimester | 4 (0.1) | 0 (0.0) | 4 (0.5) | 0 (0.0) | 0 (0.0) | 0 (0.0) | 0 (0.0) | 0 (0.0) | 1 (0.2) | 0 (0.0) | 0 (0.0) | 0 (0.0) | 0 (0.0) |
| third trimester | 2 (0.0) | 0 (0.0) | 2 (0.3) | 0 (0.0) | 0 (0.0) | 0 (0.0) | 0 (0.0) | 0 (0.0) | 1 (0.2) | 0 (0.0) | 0 (0.0) | 0 (0.0) | 0 (0.0) |
| **anti-BLyS** |  |  |  |  |  |  |  |  |  |  |  |  |  |
| prepregnancy | 2 (0.0) | 0 (0.0) | 0 (0.0) | 0 (0.0) | 0 (0.0) | 0 (0.0) | 0 (0.0) | 0 (0.0) | 2 (0.5) | 0 (0.0) | 0 (0.0) | 1 (0.6) | 0 (0.0) |
| prenatal | 3 (0.1) | 0 (0.0) | 0 (0.0) | 0 (0.0) | 0 (0.0) | 0 (0.0) | 0 (0.0) | 0 (0.0) | 3 (0.7) | 0 (0.0) | 0 (0.0) | 2 (1.1) | 0 (0.0) |
| first trimester | 3 (0.1) | 0 (0.0) | 0 (0.0) | 0 (0.0) | 0 (0.0) | 0 (0.0) | 0 (0.0) | 0 (0.0) | 3 (0.7) | 0 (0.0) | 0 (0.0) | 2 (1.1) | 0 (0.0) |
| second trimester | 1 (0.0) | 0 (0.0) | 0 (0.0) | 0 (0.0) | 0 (0.0) | 0 (0.0) | 0 (0.0) | 0 (0.0) | 1 (0.2) | 0 (0.0) | 0 (0.0) | 0 (0.0) | 0 (0.0) |
| third trimester | 0 (0.0) | 0 (0.0) | 0 (0.0) | 0 (0.0) | 0 (0.0) | 0 (0.0) | 0 (0.0) | 0 (0.0) | 0 (0.0) | 0 (0.0) | 0 (0.0) | 0 (0.0) | 0 (0.0) |
| **S1P receptor modulator** |  |  |  |  |  |  |  |  |  |  |  |  |  |
| prepregnancy | 7 (0.1) | 0 (0.0) | 0 (0.0) | 7 (1.4) | 0 (0.0) | 1 (0.1) | 0 (0.0) | 0 (0.0) | 0 (0.0) | 0 (0.0) | 0 (0.0) | 0 (0.0) | 0 (0.0) |
| prenatal | 7 (0.1) | 0 (0.0) | 0 (0.0) | 7 (1.4) | 0 (0.0) | 1 (0.1) | 0 (0.0) | 0 (0.0) | 0 (0.0) | 0 (0.0) | 0 (0.0) | 0 (0.0) | 0 (0.0) |
| first trimester | 7 (0.1) | 0 (0.0) | 0 (0.0) | 7 (1.4) | 0 (0.0) | 1 (0.1) | 0 (0.0) | 0 (0.0) | 0 (0.0) | 0 (0.0) | 0 (0.0) | 0 (0.0) | 0 (0.0) |
| second trimester | 5 (0.1) | 0 (0.0) | 0 (0.0) | 5 (1.0) | 0 (0.0) | 0 (0.0) | 0 (0.0) | 0 (0.0) | 0 (0.0) | 0 (0.0) | 0 (0.0) | 0 (0.0) | 0 (0.0) |
| third trimester | 3 (0.1) | 0 (0.0) | 0 (0.0) | 3 (0.6) | 0 (0.0) | 0 (0.0) | 0 (0.0) | 0 (0.0) | 0 (0.0) | 0 (0.0) | 0 (0.0) | 0 (0.0) | 0 (0.0) |
| **JAK inhibitor** |  |  |  |  |  |  |  |  |  |  |  |  |  |
| prepregnancy | 5 (0.1) | 1 (0.1) | 5 (0.7) | 0 (0.0) | 0 (0.0) | 0 (0.0) | 0 (0.0) | 1 (0.1) | 0 (0.0) | 0 (0.0) | 0 (0.0) | 0 (0.0) | 0 (0.0) |
| prenatal | 6 (0.1) | 1 (0.1) | 6 (0.8) | 0 (0.0) | 0 (0.0) | 0 (0.0) | 0 (0.0) | 1 (0.1) | 0 (0.0) | 0 (0.0) | 0 (0.0) | 0 (0.0) | 0 (0.0) |
| first trimester | 6 (0.1) | 1 (0.1) | 6 (0.8) | 0 (0.0) | 0 (0.0) | 0 (0.0) | 0 (0.0) | 1 (0.1) | 0 (0.0) | 0 (0.0) | 0 (0.0) | 0 (0.0) | 0 (0.0) |
| second trimester | 6 (0.1) | 1 (0.1) | 6 (0.8) | 0 (0.0) | 0 (0.0) | 0 (0.0) | 0 (0.0) | 1 (0.1) | 0 (0.0) | 0 (0.0) | 0 (0.0) | 0 (0.0) | 0 (0.0) |
| third trimester | 3 (0.1) | 1 (0.1) | 3 (0.4) | 0 (0.0) | 0 (0.0) | 0 (0.0) | 0 (0.0) | 1 (0.1) | 0 (0.0) | 0 (0.0) | 0 (0.0) | 0 (0.0) | 0 (0.0) |
| **integrin modulator** |  |  |  |  |  |  |  |  |  |  |  |  |  |
| prepregnancy | 3 (0.1) | 2 (0.2) | 1 (0.1) | 1 (0.2) | 0 (0.0) | 0 (0.0) | 0 (0.0) | 0 (0.0) | 0 (0.0) | 0 (0.0) | 0 (0.0) | 0 (0.0) | 0 (0.0) |
| prenatal | 3 (0.1) | 2 (0.2) | 1 (0.1) | 1 (0.2) | 0 (0.0) | 0 (0.0) | 0 (0.0) | 0 (0.0) | 0 (0.0) | 0 (0.0) | 0 (0.0) | 0 (0.0) | 0 (0.0) |
| first trimester | 3 (0.1) | 2 (0.2) | 1 (0.1) | 1 (0.2) | 0 (0.0) | 0 (0.0) | 0 (0.0) | 0 (0.0) | 0 (0.0) | 0 (0.0) | 0 (0.0) | 0 (0.0) | 0 (0.0) |
| second trimester | 3 (0.1) | 2 (0.2) | 1 (0.1) | 1 (0.2) | 0 (0.0) | 0 (0.0) | 0 (0.0) | 0 (0.0) | 0 (0.0) | 0 (0.0) | 0 (0.0) | 0 (0.0) | 0 (0.0) |
| third trimester | 2 (0.0) | 2 (0.2) | 1 (0.1) | 0 (0.0) | 0 (0.0) | 0 (0.0) | 0 (0.0) | 0 (0.0) | 0 (0.0) | 0 (0.0) | 0 (0.0) | 0 (0.0) | 0 (0.0) |
| **PDE4 inhibitor targeted synthetic** |  |  |  |  |  |  |  |  |  |  |  |  |  |
| prepregnancy | 3 (0.1) | 0 (0.0) | 2 (0.3) | 0 (0.0) | 2 (1.0) | 3 (0.2) | 0 (0.0) | 1 (0.1) | 0 (0.0) | 0 (0.0) | 0 (0.0) | 0 (0.0) | 1 (0.7) |
| prenatal | 3 (0.1) | 0 (0.0) | 2 (0.3) | 0 (0.0) | 2 (1.0) | 3 (0.2) | 0 (0.0) | 1 (0.1) | 0 (0.0) | 0 (0.0) | 0 (0.0) | 0 (0.0) | 1 (0.7) |
| first trimester | 3 (0.1) | 0 (0.0) | 2 (0.3) | 0 (0.0) | 2 (1.0) | 3 (0.2) | 0 (0.0) | 1 (0.1) | 0 (0.0) | 0 (0.0) | 0 (0.0) | 0 (0.0) | 1 (0.7) |
| second trimester | 3 (0.1) | 0 (0.0) | 2 (0.3) | 0 (0.0) | 2 (1.0) | 3 (0.2) | 0 (0.0) | 1 (0.1) | 0 (0.0) | 0 (0.0) | 0 (0.0) | 0 (0.0) | 1 (0.7) |
| third trimester | 2 (0.0) | 0 (0.0) | 1 (0.1) | 0 (0.0) | 2 (1.0) | 2 (0.1) | 0 (0.0) | 0 (0.0) | 0 (0.0) | 0 (0.0) | 0 (0.0) | 0 (0.0) | 0 (0.0) |
| **anti-cd20** |  |  |  |  |  |  |  |  |  |  |  |  |  |
| prepregnancy | 1 (0.0) | 0 (0.0) | 0 (0.0) | 1 (0.2) | 0 (0.0) | 0 (0.0) | 0 (0.0) | 0 (0.0) | 0 (0.0) | 0 (0.0) | 0 (0.0) | 0 (0.0) | 0 (0.0) |
| prenatal | 2 (0.0) | 0 (0.0) | 0 (0.0) | 2 (0.4) | 0 (0.0) | 0 (0.0) | 0 (0.0) | 0 (0.0) | 0 (0.0) | 0 (0.0) | 0 (0.0) | 0 (0.0) | 0 (0.0) |
| first trimester | 1 (0.0) | 0 (0.0) | 0 (0.0) | 1 (0.2) | 0 (0.0) | 0 (0.0) | 0 (0.0) | 0 (0.0) | 0 (0.0) | 0 (0.0) | 0 (0.0) | 0 (0.0) | 0 (0.0) |
| second trimester | 1 (0.0) | 0 (0.0) | 0 (0.0) | 1 (0.2) | 0 (0.0) | 0 (0.0) | 0 (0.0) | 0 (0.0) | 0 (0.0) | 0 (0.0) | 0 (0.0) | 0 (0.0) | 0 (0.0) |
| third trimester | 2 (0.0) | 0 (0.0) | 0 (0.0) | 2 (0.4) | 0 (0.0) | 0 (0.0) | 0 (0.0) | 0 (0.0) | 0 (0.0) | 0 (0.0) | 0 (0.0) | 0 (0.0) | 0 (0.0) |
| **anti-cd52** |  |  |  |  |  |  |  |  |  |  |  |  |  |
| prepregnancy | 0 (0.0) | 0 (0.0) | 0 (0.0) | 0 (0.0) | 0 (0.0) | 0 (0.0) | 0 (0.0) | 0 (0.0) | 0 (0.0) | 0 (0.0) | 0 (0.0) | 0 (0.0) | 0 (0.0) |
| prenatal | 0 (0.0) | 0 (0.0) | 0 (0.0) | 0 (0.0) | 0 (0.0) | 0 (0.0) | 0 (0.0) | 0 (0.0) | 0 (0.0) | 0 (0.0) | 0 (0.0) | 0 (0.0) | 0 (0.0) |
| first trimester | 0 (0.0) | 0 (0.0) | 0 (0.0) | 0 (0.0) | 0 (0.0) | 0 (0.0) | 0 (0.0) | 0 (0.0) | 0 (0.0) | 0 (0.0) | 0 (0.0) | 0 (0.0) | 0 (0.0) |
| second trimester | 0 (0.0) | 0 (0.0) | 0 (0.0) | 0 (0.0) | 0 (0.0) | 0 (0.0) | 0 (0.0) | 0 (0.0) | 0 (0.0) | 0 (0.0) | 0 (0.0) | 0 (0.0) | 0 (0.0) |
| third trimester | 0 (0.0) | 0 (0.0) | 0 (0.0) | 0 (0.0) | 0 (0.0) | 0 (0.0) | 0 (0.0) | 0 (0.0) | 0 (0.0) | 0 (0.0) | 0 (0.0) | 0 (0.0) | 0 (0.0) |
| **budesonide** |  |  |  |  |  |  |  |  |  |  |  |  |  |
| prepregnancy | 10 (0.2) | 10 (0.8) | 0 (0.0) | 0 (0.0) | 0 (0.0) | 0 (0.0) | 0 (0.0) | 0 (0.0) | 0 (0.0) | 0 (0.0) | 0 (0.0) | 0 (0.0) | 0 (0.0) |
| prenatal | 22 (0.4) | 22 (1.7) | 1 (0.1) | 0 (0.0) | 0 (0.0) | 0 (0.0) | 0 (0.0) | 0 (0.0) | 0 (0.0) | 0 (0.0) | 0 (0.0) | 0 (0.0) | 0 (0.0) |
| first trimester | 14 (0.2) | 14 (1.1) | 0 (0.0) | 0 (0.0) | 0 (0.0) | 0 (0.0) | 0 (0.0) | 0 (0.0) | 0 (0.0) | 0 (0.0) | 0 (0.0) | 0 (0.0) | 0 (0.0) |
| second trimester | 14 (0.2) | 14 (1.1) | 1 (0.1) | 0 (0.0) | 0 (0.0) | 0 (0.0) | 0 (0.0) | 0 (0.0) | 0 (0.0) | 0 (0.0) | 0 (0.0) | 0 (0.0) | 0 (0.0) |
| third trimester | 15 (0.3) | 15 (1.2) | 1 (0.1) | 0 (0.0) | 0 (0.0) | 0 (0.0) | 0 (0.0) | 0 (0.0) | 0 (0.0) | 0 (0.0) | 0 (0.0) | 0 (0.0) | 0 (0.0) |
| **steroids** |  |  |  |  |  |  |  |  |  |  |  |  |  |
| prepregnancy | 108 (1.9) | 37 (2.9) | 27 (3.7) | 2 (0.4) | 4 (1.9) | 13 (0.7) | 1 (1.9) | 10 (1.5) | 22 (5.0) | 2 (2.4) | 3 (5.3) | 5 (2.8) | 3 (2.0) |
| prenatal | 479 (8.3) | 154 (11.9) | 108 (14.8) | 28 (5.5) | 18 (8.7) | 79 (4.2) | 2 (3.7) | 38 (5.6) | 73 (16.6) | 13 (15.5) | 9 (15.8) | 18 (10.0) | 13 (8.8) |
| first trimester | 208 (3.6) | 77 (6.0) | 51 (7.0) | 11 (2.2) | 8 (3.9) | 24 (1.3) | 1 (1.9) | 20 (3.0) | 29 (6.6) | 4 (4.8) | 3 (5.3) | 9 (5.0) | 4 (2.7) |
| second trimester | 205 (3.5) | 67 (5.2) | 48 (6.6) | 8 (1.6) | 7 (3.4) | 31 (1.7) | 1 (1.9) | 18 (2.7) | 32 (7.3) | 7 (8.3) | 4 (7.0) | 6 (3.3) | 5 (3.4) |
| third trimester | 304 (5.3) | 99 (7.7) | 68 (9.3) | 15 (3.0) | 12 (5.8) | 52 (2.8) | 2 (3.7) | 23 (3.4) | 45 (10.3) | 9 (10.7) | 7 (12.3) | 10 (5.6) | 8 (5.4) |

GA Gestational Age; LMP Last Menstrual Period; IMMs Immunomodulatory Medication;

RxNorm codes of IMMs are listed in Table S3. Prepregnancy exposure indicates exposure status during 180 days prepregnancy period (LMP-180days~LMP). Prenatal exposure indicates exposure status from LMP to delivery. First trimester exposure indicates exposure status from LMP to end of first trimester (LMP≤exposure<GA 13 weeks). Second trimester exposure indicates exposure status from end of first trimester to end of second trimester (GA 13 weeks≤exposure<GA 28 weeks). Third trimester exposure indicates exposure status from end of second trimester to delivery date (GA 28 weeks≤exposure<date of delivery)

#### **Table S5 Statistical significance of descriptive statistics**

| **variable** | **IMIDs** | **IBD** | **RA** | **MS** | **PsA** | **Ps** | **SSc** | **SpA** | **SLE** | **Va** | **Sc** | **APS** | **SjS** |
| --- | --- | --- | --- | --- | --- | --- | --- | --- | --- | --- | --- | --- | --- |
| Maternal age | **** | **** | **** | **** | **** | **** | * | **** | **** | ns | ** | **** | **** |
| Age group | **** | **** | * | **** | *** | **** | ns | **** | *** | ns | ns | + | ** |
| Race group | **** | **** | *** | **** | ** | **** | ns | **** | * | ns | ns | + | ns |
| Ethnicity | **** | **** | * | **** | **** | **** | ns | **** | ns | ns | ns | + | ns |
| BMI category | **** | **** | **** | ns | + | **** | ns | **** | + | ns | ns | ns | ns |
| insurance | **** | **** | * | **** | ns | **** | ns | ns | ns | ns | ns | ns | ** |
| smoker | **** | ns | + | * | ns | *** | ns | ** | ns | ns | ns | ns | ns |
| Illegal drug user | **** | **** | * | + | ns | * | ns | **** | * | ns | ns | ns | ns |
| Alcohol user | **** | **** | **** | **** | ** | **** | ** | *** | **** | ns | *** | ns | ** |
| Vulnerability index of socioeconomic status | **** | **** | *** | **** | ** | **** | ns | ns | ns | ns | ns | ns | ns |
| Vulnerability index of housing composition | *** | ** | ns | ns | + | **** | ns | ** | ns | ns | ns | ns | ns |
| Vulnerability index of minority status and language | **** | **** | **** | **** | ** | **** | ns | **** | ns | ns | * | ns | ns |
| Vulnerability index of housing type and transportation | *** | **** | ns | ns | * | ns | ns | ns | ns | ns | ns | ns | ns |
| Rural/urban classification | **** | ** | + | ns | ns | **** | ns | * | ns | ns | ns | ns | ns |
| Gravidity | **** | ns | ns | * | * | ** | ns | ** | * | ns | ** | **** | ns |
| Parity | ** | + | ns | ** | ns | ns | ns | ns | ns | ns | ** | * | ns |
| Preterm_history | *** | + | ns | ** | ns | * | ns | ns | * | ns | ** | *** | ns |
| Fetal sex | ns | ns | ns | **** | ns | ns | ns | ns | ** | ns | ns | ns | ns |
| Delivery year | **** | **** | **** | ns | *** | **** | ns | **** | **** | ns | ns | ** | ** |
| Diabetes | **** | ns | *** | ns | * | **** | ns | + | ns | ns | ns | ns | ns |
| Chronic kidney disease | **** | + | ns | ns | ns | ns | ns | ns | **** | * | ns | * | ns |
| Obesity | **** | ns | **** | + | **** | **** | ns | **** | **** | ns | ** | ** | ** |
| Asthma | **** | **** | **** | *** | **** | **** | ns | **** | **** | * | ns | **** | **** |
| Chronic lung disease | ns | ns | ns | ns | ns | ns | ns | ns | ns | ns | ns | ns | ns |
| Depression | **** | **** | **** | **** | **** | **** | *** | **** | **** | * | + | **** | **** |
| Pneumonia | **** | ns | ** | + | ** | ** | ns | ** | *** | * | **** | **** | ns |
| Urinary tract infection | **** | **** | **** | **** | *** | **** | ns | **** | **** | **** | * | **** | ns |
| Sexually transmitted disease | **** | **** | ** | ** | ns | **** | ns | **** | ns | ns | ns | * | * |
| Cardiovascular disease | **** | **** | **** | **** | **** | **** | **** | **** | **** | **** | **** | **** | **** |
| Sepsis | **** | **** | ** | ns | * | ns | ns | *** | **** | ns | ns | ns | ns |

APS Antiphospholipid Syndrome; IBD Inflammatory Bowel Disease; IMID Immune-mediated Inflammatory Disease; MS Multiple Sclerosis; Ps Psoriasis; PsA Psoriatic Arthritis; Sc Sarcoidosis; SLE Systemic Lupus Erythematosus; SpA Spondyloarthritis; SjS Sjörgen’s Syndrome; SSc Systemic Sclerosis; Va Vasculitis;

Variables are defined in Table S2. SNOMED codes of diagnoses are listed in Table S1. Difference in distribution of corresponding variable between IMIDs and control was evaluated using t-test, fisher exact test, and chi-square test for continuous, binary, and categorical variables. Multiple testing error was corrected using Benjamin-Hochberg Statistical significance was reported as follows. p<0·0001:****, 0·0001≤p<0·001:***, 0·001≤p<0·01:**, 0·01≤p<0·05:*, 0·05≤p<0·1:+, 0.1<p:ns

#### **Table S6. Standardized mean differences after propensity score matching for individual IMIDs group**

| **variable** | **IMIDs** | **IBD** | **RA** | **MS** | **PsA** | **Ps** | **SSc** | **SpA** | **SLE** | **Va** | **Sc** | **APS** | **SjS** |
| --- | --- | --- | --- | --- | --- | --- | --- | --- | --- | --- | --- | --- | --- |
| Pregravid BMI | 0.05 | 0.05 | -0.02 | 0.03 | 0.09 | 0.01 | 0.19 | -0.02 | 0.04 | -0.03 | 0.02 | 0.16 | -0.04 |
| Maternal age | 0.02 | 0.02 | -0.04 | -0.08 | 0.00 | -0.04 | 0.02 | -0.06 | -0.04 | 0.07 | 0.00 | 0.05 | -0.02 |
| Ethnic group | 0.04 | 0.04 | 0.03 | 0.06 | -0.14 | 0.02 | -0.04 | 0.01 | -0.02 | 0.00 | 0.00 | -0.06 | 0.13 |
| Fetal sex | -0.00 | -0.00 | -0.01 | 0.01 | 0.04 | 0.03 | -0.13 | -0.05 | -0.05 | 0.02 | 0.03 | -0.16 | -0.04 |
| Parity | -0.02 | -0.02 | -0.04 | -0.01 | 0.06 | -0.04 | -0.16 | -0.03 | 0.03 | 0.03 | -0.07 | -0.05 | 0.17 |
| Gravidity | 0.00 | 0.00 | -0.03 | -0.01 | 0.03 | -0.02 | -0.03 | 0.07 | -0.04 | -0.09 | -0.03 | 0.04 | 0.05 |
| Preterm history | 0.01 | 0.01 | 0.01 | -0.03 | -0.03 | 0.04 | -0.11 | 0.04 | 0.00 | 0.07 | -0.22 | 0.06 | -0.06 |
| Illegal drug user | 0.02 | 0.02 | 0.08 | -0.02 | 0.02 | 0.01 | -0.16 | -0.01 | 0.04 | 0.00 | -0.11 | -0.08 | -0.03 |
| smoker | -0.02 | -0.02 | 0.05 | 0.02 | 0.12 | 0.01 | -0.12 | 0.01 | 0.09 | -0.04 | 0.00 | 0.04 | -0.03 |
| Vulnerability index of socioeconomic status | 0.00 | 0.00 | 0.06 | -0.00 | 0.08 | 0.04 | -0.16 | 0.07 | 0.02 | 0.01 | 0.05 | -0.14 | -0.08 |
| Vulnerability index of housing composition | -0.01 | -0.01 | 0.02 | -0.02 | 0.12 | -0.01 | 0.00 | 0.05 | 0.01 | -0.07 | 0.00 | 0.04 | 0.12 |
| Vulnerability index of minority status and language | 0.02 | 0.02 | 0.04 | -0.02 | 0.09 | 0.03 | -0.13 | 0.05 | 0.03 | 0.07 | 0.13 | -0.06 | -0.12 |
| Vulnerability index of housing type and transportation | 0.02 | 0.02 | 0.01 | -0.08 | 0.15 | 0.02 | -0.18 | 0.06 | 0.03 | 0.11 | 0.04 | -0.17 | -0.16 |
| Rural/urban classification | 0.04 | 0.04 | -0.08 | -0.04 | -0.06 | -0.05 | 0.13 | -0.03 | -0.08 | -0.15 | -0.02 | -0.06 | -0.01 |
| Diabetes | 0.04 | 0.04 | 0.05 | -0.02 | -0.08 | 0.00 | NaN | -0.04 | 0.00 | -0.09 | -0.11 | 0.11 | 0.12 |
| Chronic kidney disease | 0.02 | 0.02 | 0.05 | NaN | NaN | 0.03 | NaN | NaN | 0.05 | 0.00 | NaN | 0.06 | 0.00 |
| Obesity | 0.03 | 0.03 | 0.06 | 0.02 | 0.06 | 0.09 | NaN | -0.05 | 0.02 | 0.05 | 0.18 | 0.11 | 0.08 |
| Chronic liver disease | 0.01 | 0.01 | -0.03 | -0.06 | NaN | -0.05 | 0.00 | -0.02 | 0.07 | -0.09 | NaN | NaN | 0.00 |
| Asthma | 0.01 | 0.01 | 0.03 | -0.04 | 0.01 | 0.03 | -0.12 | 0.03 | 0.01 | 0.11 | -0.15 | 0.08 | 0.09 |
| Depression | 0.04 | 0.04 | 0.02 | 0.06 | -0.04 | 0.01 | -0.24 | 0.07 | -0.06 | -0.09 | 0.00 | -0.01 | -0.12 |
| Pneumonia | 0.04 | 0.04 | 0.03 | 0.00 | 0.03 | 0.05 | NaN | 0.05 | 0.00 | -0.05 | 0.00 | 0.07 | -0.12 |
| Urinary tract infection | 0.02 | 0.02 | 0.01 | 0.04 | -0.04 | -0.03 | 0.17 | 0.00 | 0.02 | 0.13 | -0.08 | 0.11 | 0.08 |
| Sexually transmitted disease | -0.00 | -0.00 | -0.09 | -0.06 | 0.06 | 0.01 | 0.09 | 0.04 | -0.01 | 0.07 | -0.11 | 0.00 | 0.10 |
| Cardiovascular disease | 0.02 | 0.02 | -0.00 | 0.05 | -0.05 | 0.01 | -0.15 | 0.01 | 0.00 | 0.00 | 0.07 | -0.04 | -0.04 |
| Sepsis | 0.03 | 0.03 | 0.00 | -0.02 | 0.04 | -0.03 | 0.27 | 0.03 | -0.01 | -0.07 | NaN | NaN | NaN |
| Year_2013 | -0.01 | -0.01 | -0.05 | -0.02 | -0.03 | 0.00 | 0.19 | 0.05 | 0.06 | 0.15 | -0.18 | 0.00 | -0.04 |
| Year_2014 | 0.03 | 0.03 | 0.01 | 0.05 | -0.08 | -0.03 | 0.20 | 0.03 | 0.03 | 0.00 | 0.00 | 0.09 | 0.10 |
| Year_2015 | -0.01 | -0.01 | -0.03 | 0.02 | 0.00 | 0.01 | -0.07 | 0.05 | -0.01 | -0.05 | 0.16 | 0.05 | 0.03 |
| Year_2016 | -0.02 | -0.02 | -0.06 | 0.00 | -0.11 | 0.02 | 0.07 | 0.04 | -0.01 | 0.06 | -0.13 | 0.00 | -0.03 |
| Year_2017 | -0.06 | -0.06 | 0.08 | -0.10 | 0.00 | -0.02 | -0.07 | -0.04 | 0.03 | 0.26 | 0.16 | 0.02 | -0.04 |
| Year_2018 | 0.02 | 0.02 | 0.00 | -0.03 | -0.04 | 0.03 | -0.06 | -0.06 | 0.00 | -0.10 | -0.15 | 0.02 | -0.14 |
| Year_2019 | -0.01 | -0.01 | 0.04 | 0.05 | -0.12 | -0.03 | 0.12 | -0.04 | 0.02 | 0.00 | 0.18 | -0.09 | 0.08 |
| Year_2020 | 0.00 | 0.00 | -0.01 | 0.05 | 0.14 | 0.00 | -0.09 | 0.03 | -0.01 | 0.03 | 0.06 | 0.07 | 0.04 |
| Year_2021 | 0.01 | 0.01 | 0.02 | 0.00 | 0.05 | -0.03 | -0.05 | 0.01 | 0.01 | 0.04 | 0.16 | -0.07 | -0.04 |
| Year_2022 | 0.04 | 0.04 | -0.01 | -0.01 | 0.12 | 0.05 | 0.00 | 0.00 | -0.08 | -0.17 | -0.11 | 0.00 | 0.08 |
| Race_white | -0.09 | -0.09 | -0.01 | -0.02 | -0.10 | -0.05 | -0.21 | -0.01 | -0.08 | 0.03 | 0.11 | -0.06 | -0.01 |
| Race_asian | 0.04 | 0.04 | -0.01 | 0.00 | 0.11 | -0.02 | -0.09 | -0.01 | 0.02 | -0.07 | 0.00 | 0.05 | -0.13 |
| Race_black | 0.05 | 0.05 | 0.04 | 0.00 | 0.06 | 0.05 | NaN | 0.02 | 0.08 | 0.13 | -0.08 | -0.07 | -0.11 |
| Race_other | 0.03 | 0.03 | 0.03 | 0.02 | 0.02 | 0.04 | 0.16 | 0.03 | 0.01 | -0.13 | -0.09 | 0.08 | 0.19 |
| Commercial insurance | -0.01 | -0.01 | -0.04 | -0.07 | 0.02 | -0.04 | -0.07 | -0.01 | 0.01 | -0.02 | 0.03 | 0.00 | -0.07 |

APS Antiphospholipid Syndrome; IBD Inflammatory Bowel Disease; IMID Immune-mediated Inflammatory Disease; MS Multiple Sclerosis; Ps Psoriasis; PsA Psoriatic Arthritis; Sc Sarcoidosis; SLE Systemic Lupus Erythematosus; SpA Spondyloarthritis; SjS Sjörgen’s Syndrome; SSc Systemic Sclerosis; Va Vasculitis; IMM Immunomodulatory Medication; LMP Last Menstrual Period; GA Gestational Age

The standardized mean difference below 0.2 is considered a small effect size. ^3^

#### **Table S7 Standardized mean difference after propensity score matching for individual IMIDs group in the sensitivity analysis**

| **variable** | **IMIDs** | **IBD** | **RA** | **MS** | **PsA** | **Ps** | **SSc** | **SpA** | **SLE** | **Va** | **Sc** | **APS** | **SjS** |
| --- | --- | --- | --- | --- | --- | --- | --- | --- | --- | --- | --- | --- | --- |
| Pregravid BMI | 0.03 | -0.01 | 0.03 | 0.03 | 0.11 | 0.03 | -0.15 | 0.06 | -0.02 | -0.13 | -0.06 | -0.09 | 0.06 |
| Maternal age | -0.02 | 0.01 | -0.04 | -0.08 | -0.11 | 0.00 | -0.07 | -0.00 | 0.03 | -0.07 | 0.23 | -0.14 | -0.03 |
| Ethnic group | -0.01 | 0.04 | 0.01 | -0.02 | -0.07 | 0.00 | 0.04 | -0.02 | 0.01 | -0.03 | 0.11 | 0.01 | -0.05 |
| Fetal sex | 0.00 | 0.01 | 0.00 | 0.07 | 0.00 | -0.01 | 0.00 | 0.03 | -0.08 | 0.02 | 0.11 | 0.00 | 0.00 |
| Parity | 0.01 | -0.04 | -0.01 | 0.00 | -0.04 | 0.06 | -0.14 | 0.09 | -0.07 | 0.16 | 0.03 | 0.00 | 0.03 |
| Gravidity | 0.03 | 0.01 | 0.03 | 0.03 | -0.08 | 0.08 | 0.03 | 0.09 | -0.05 | 0.02 | 0.11 | 0.07 | 0.05 |
| Preterm history | 0.00 | 0.03 | -0.01 | 0.06 | 0.10 | 0.01 | -0.11 | 0.04 | -0.05 | 0.00 | 0.11 | 0.00 | -0.06 |
| Illegal drug user | 0.05 | 0.03 | 0.03 | 0.03 | 0.05 | 0.05 | 0.27 | 0.04 | -0.01 | 0.03 | 0.12 | -0.08 | 0.11 |
| Smoker | 0.03 | 0.02 | 0.00 | 0.14 | 0.12 | 0.04 | 0.00 | 0.03 | -0.04 | -0.10 | 0.00 | -0.09 | 0.03 |
| Vulnerability index of socioeconomic status | 0.03 | -0.00 | 0.02 | -0.00 | 0.24 | 0.06 | -0.17 | 0.04 | 0.01 | 0.08 | 0.12 | 0.02 | 0.07 |
| Vulnerability index of housing composition | 0.05 | 0.01 | 0.02 | 0.05 | 0.18 | 0.02 | 0.05 | 0.01 | -0.05 | 0.13 | 0.06 | -0.09 | 0.13 |
| Vulnerability index of minority status and language | 0.01 | 0.01 | 0.01 | -0.07 | 0.07 | 0.02 | -0.06 | 0.05 | -0.03 | -0.12 | 0.10 | -0.02 | 0.12 |
| Vulnerability index of housing type and transportation | 0.01 | -0.03 | 0.02 | -0.00 | 0.09 | 0.04 | -0.06 | 0.01 | -0.02 | 0.16 | -0.02 | 0.03 | 0.12 |
| Rural/urban classification | -0.06 | -0.01 | -0.01 | 0.09 | -0.06 | -0.05 | -0.07 | -0.03 | -0.09 | -0.11 | -0.17 | -0.13 | -0.07 |
| Year_2013 | -0.00 | -0.02 | -0.01 | -0.02 | -0.05 | 0.00 | 0.19 | -0.01 | 0.06 | 0.00 | -0.06 | 0.06 | -0.07 |
| Year_2014 | -0.01 | -0.01 | -0.03 | 0.00 | 0.02 | -0.02 | -0.07 | -0.03 | 0.02 | 0.05 | -0.06 | 0.00 | 0.10 |
| Year_2015 | 0.00 | -0.04 | 0.02 | -0.01 | 0.07 | -0.02 | 0.20 | -0.02 | 0.02 | 0.00 | 0.00 | -0.02 | 0.08 |
| Year_2016 | -0.01 | 0.03 | 0.04 | -0.01 | -0.07 | 0.00 | -0.13 | 0.04 | 0.02 | -0.18 | 0.00 | -0.17 | -0.03 |
| Year_2017 | -0.02 | -0.03 | 0.04 | 0.08 | -0.08 | 0.01 | 0.00 | 0.00 | -0.09 | 0.04 | 0.39 | -0.04 | -0.08 |
| Year_2018 | 0.00 | 0.03 | -0.00 | -0.01 | -0.08 | 0.03 | 0.22 | 0.01 | 0.02 | -0.07 | -0.05 | 0.00 | 0.02 |
| Year_2019 | 0.00 | 0.02 | 0.00 | -0.04 | -0.06 | -0.03 | 0.00 | 0.01 | -0.01 | 0.00 | -0.10 | -0.03 | -0.07 |
| Year_2020 | -0.00 | -0.03 | -0.05 | -0.01 | 0.05 | -0.01 | -0.18 | 0.10 | -0.02 | 0.10 | -0.05 | -0.05 | -0.02 |
| Year_2021 | 0.01 | -0.02 | -0.01 | -0.01 | 0.19 | -0.01 | 0.10 | -0.04 | 0.08 | 0.00 | 0.07 | 0.20 | 0.02 |
| Year_2022 | 0.02 | 0.04 | 0.01 | 0.03 | 0.01 | 0.04 | -0.05 | -0.05 | -0.05 | 0.03 | 0.06 | 0.06 | 0.06 |
| Race_white | -0.02 | -0.02 | 0.01 | 0.03 | 0.00 | -0.06 | 0.12 | -0.08 | 0.07 | -0.09 | -0.15 | -0.05 | -0.03 |
| Race_asian | 0.01 | -0.01 | -0.05 | -0.04 | 0.04 | 0.01 | -0.16 | 0.01 | -0.05 | -0.07 | 0.00 | 0.02 | 0.04 |
| Race_black | -0.01 | 0.00 | -0.01 | 0.04 | -0.08 | -0.01 | NaN | 0.02 | -0.09 | 0.13 | -0.16 | 0.00 | 0.04 |
| Race_other | -0.01 | 0.03 | -0.04 | 0.02 | -0.09 | 0.03 | -0.05 | 0.02 | 0.03 | 0.05 | 0.10 | 0.06 | -0.04 |
| Commercial insurance | -0.05 | -0.01 | -0.03 | -0.04 | -0.10 | -0.06 | -0.23 | -0.03 | 0.08 | -0.14 | 0.14 | -0.08 | -0.03 |

APS Antiphospholipid Syndrome; IBD Inflammatory Bowel Disease; IMID Immune-mediated Inflammatory Disease; MS Multiple Sclerosis; Ps Psoriasis; PsA Psoriatic Arthritis; Sc Sarcoidosis; SLE Systemic Lupus Erythematosus; SpA Spondyloarthritis; SjS Sjörgen’s Syndrome; SSc Systemic Sclerosis; Va Vasculitis; IMMs Immunomodulatory Medications; LMP Last Menstrual Period; GA Gestational Age

The standardized mean difference below 0.2 is considered a small effect size. ^3^

#### **Table S8 Full list of packages used and their version number**

| Package name | Version number |
| --- | --- |
| imbalanced-learn | 0.10.1 |
| ipython | 8.5.0 |
| joblib | 1.1.1 |
| joblibspark | 0.5.1 |
| matplotlib | 3.5.1 |
| matplotlib-inline | 0.1.2 |
| numpy | 1.21.5 |
| pandas | 1.4.2 |
| pip | 21.2.4 |
| scikit-learn | 1.0.2 |
| scipy | 1.7.3 |
| seaborn | 0.11.2 |
| statsmodels | 0.13.2 |

### **Supplementary Figures**

#### **Figure S1. Prenatal IMM prescription rate of individual IMIDs**

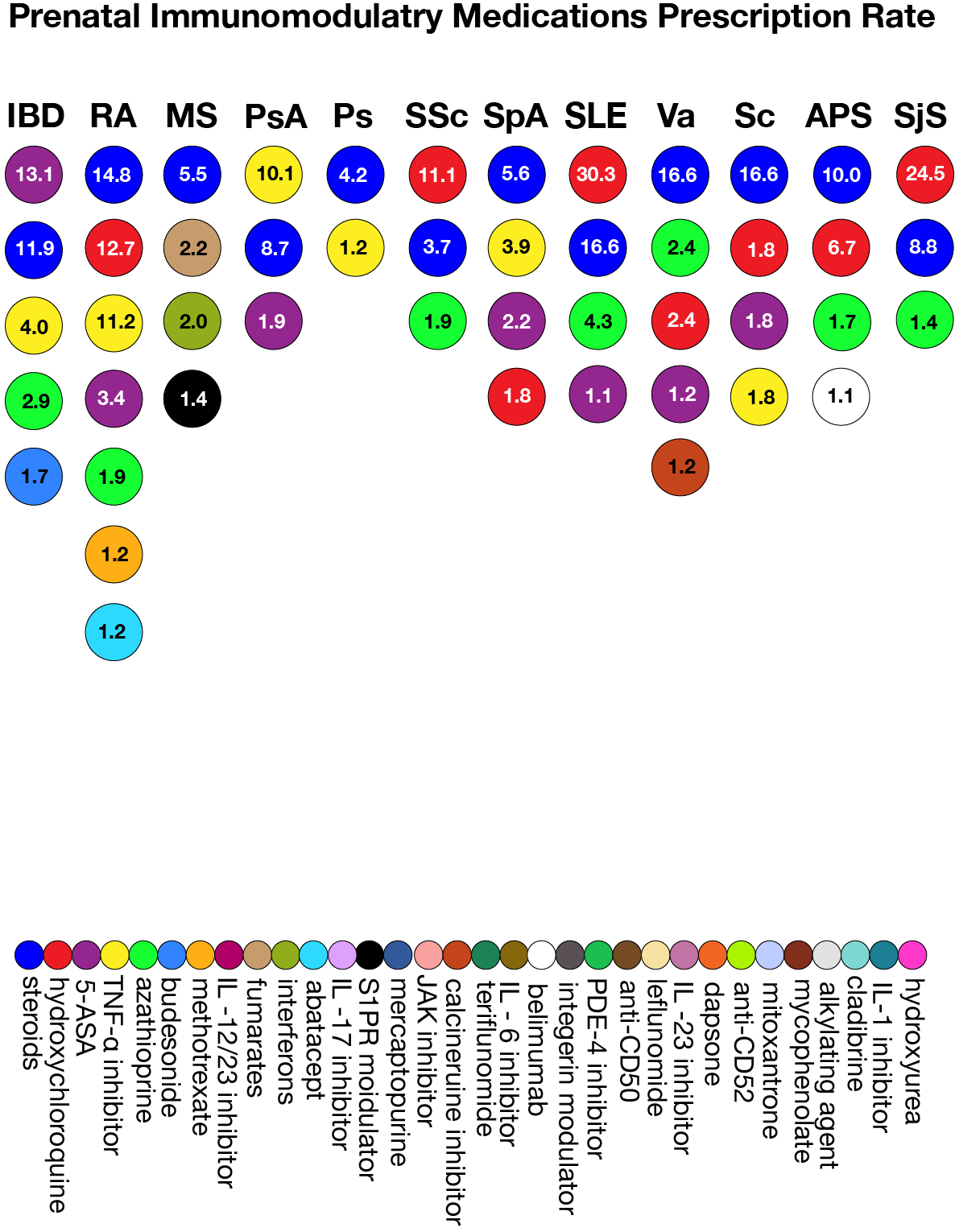

APS Antiphospholipid Syndrome; IBD Inflammatory Bowel Disease; IMIDs Immune-mediated Inflammatory Disease; MS Multiple Sclerosis; Ps Psoriasis; PsA Psoriatic Arthritis; Sc Sarcoidosis; SLE Systemic Lupus Erythematosus; SpA Spondyloarthritis; SjS Sjörgen’s Syndrome; SSc Systemic Sclerosis; Va Vasculitis;

RxNorm codes of immunomodulatry medications (IMMs) are listed in the Table S3. Each column indicates ranking of individual IMID’s prenatal IMMs prescription rate. Prenatal IMMs prescription rate are displayed in the descending order. IMMs with prescription rate below 1% are not displayed. IMMs prescription rate widely varied depending on the individual IMID.

#### **Figure S2. Standardized mean difference before and after propensity score matching for the IMIDs group**

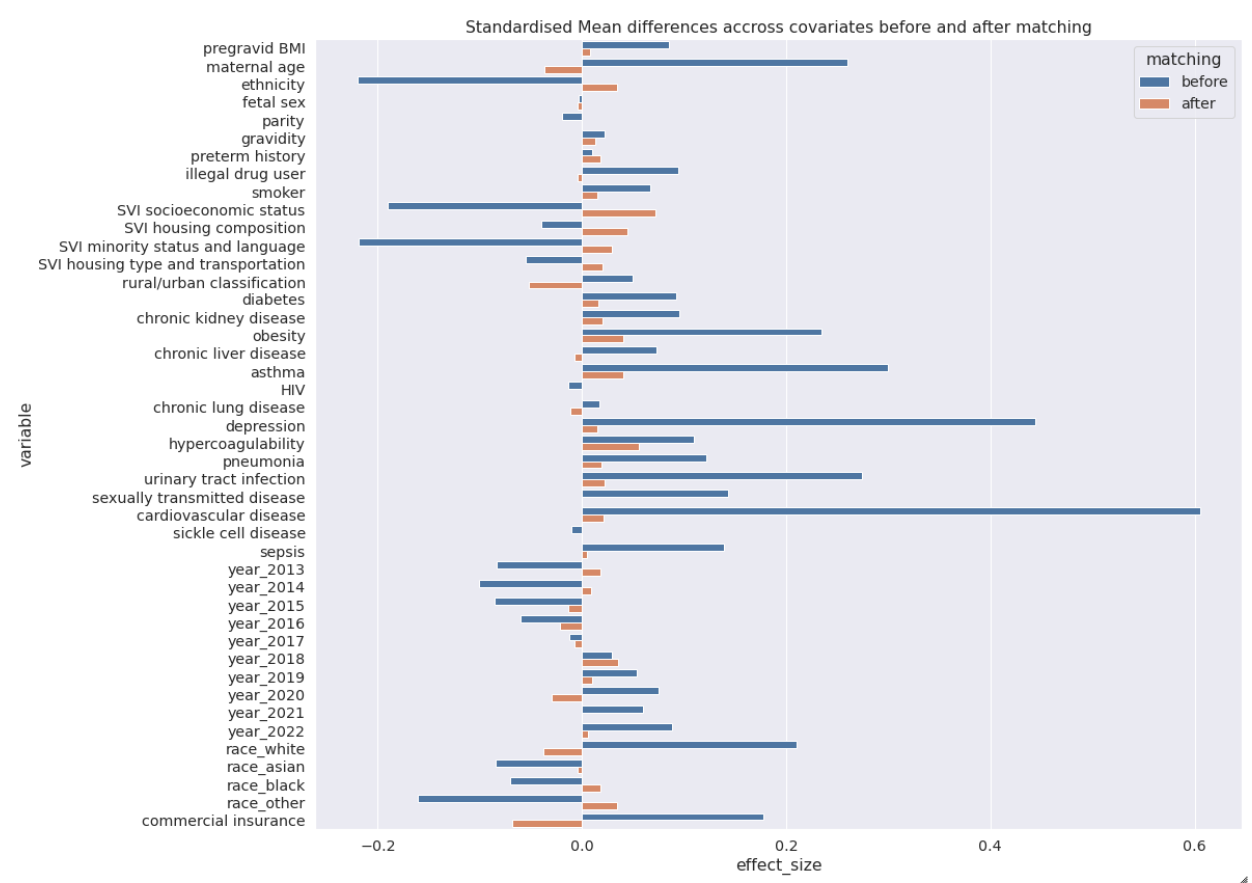

Variables are defined in Table S1. The standardized mean difference after propensity score matching for individual IMIDs groups is shown in Table S6. The standardized mean difference of sensitivity analysis is shown in Table S7.

#### **Figure S3. Hydroxychloroquine usage pattern from 2020 to 2022**

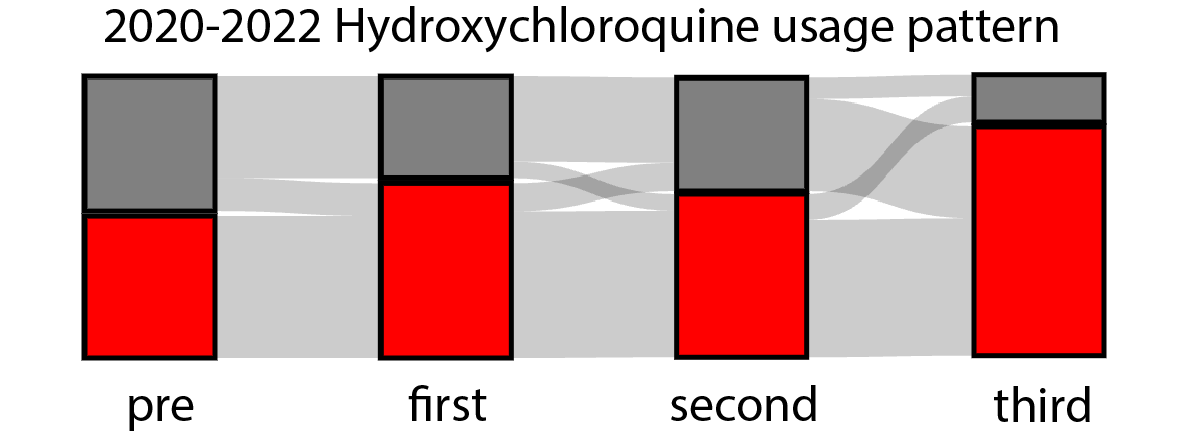

Hydroxychloroquine prescription patterns among patients who prescribed at least once from LMP-180 days to delivery date in 2020-2022. Pre, first, second, and third columns indicate 180 days prepregnancy period, first second and third trimester. Red and gray portions respectivley indicate exposed and unexposed patients for corresponding time periods.
